# Clofazimine treatment modulates key non-coding RNAs associated with tumor progression and drug resistance in lethal Prostate Cancer

**DOI:** 10.1101/2025.02.20.25322381

**Authors:** Sarah Batten, Harish Kumar, Jeremiah Pfitzer, Daniel Chinedu Nweze, Suman Mazumder, Robert Arnold, Panagiotis Mistriotis, Taraswi Mitra Ghosh, Amit Kumar Mitra

## Abstract

Prostate cancer (PCa) is the most commonly diagnosed cancer and the second-leading cause of cancer death among men in the United States, representing 24.3% of all new cancer cases in the US. Metastatic castration-resistant prostate cancer (mCRPC) is a clinically advanced form of PCa that is associated with increased aggressiveness, cancer stemness, morbidity, and the risk of developing resistance to taxanes, currently the first-line chemotherapeutic agents for mCRPC.Development of new target-directed drugs to treat mCRPC poses significant challenges, given the recognition that monospecific inhibitors have limited efficacy. Hence, there is an urgent medical need to develop new strategies that block major oncogenic signaling pathways driving the most lethal forms of PCa.

Clofazimine (CLF) is a potential immunomodulator drug that is FDA-approved for the treatment of leprosy. Recently, using a phenotype-based high-throughput drug screening, we demonstrated the *in vitro* (cell lines), *in vivo* (mouse xenograft models), and *ex vivo* (patient-derived primary tumor cells) efficacy of CLF in drug-resistant forms of chronic myeloid leukemia and multiple myeloma.

In this study, we demonstrate that CLF is effective as a single agent and in combination with docetaxel (DTX) in a panel of PCa cell lines representing the diversity of CRPC patients. We also found that CLF reduces aldehyde dehydrogenase activity, which is a marker for cancer ‘stem-like’ cells (CSCs), a subtype of cancer cells with self-renewal and differentiation capacities (acquisition of mesenchymal phenotype or epithelial to mesenchymal transdifferentiation/EMT) that significantly contribute to tumor aggressiveness and the development of drug resistance. Further, using a microfluidic assay, we showed the impact of CLF on cancer cell migration and metastatic potential. Drug-induced changes were investigated using bulk tumor and single-cell RNA sequencing followed by functional analysis of top gene/pathway signatures, where CLF treatment was found to modulate cellular pathways associated with apoptosis, ER stress, oxidative phosphorylation, and mitochondrial dysfunction. Most importantly, CLF modulates the expression of several non-coding RNAs, including MALAT1 and NEAT1, that are linked to tumor cell proliferation, cell migration, and drug resistance. *In silico* validation of the non-coding RNA signature was performed using multiple patient datasets.

Our results support the preclinical development of CLF against lethal PCa and provide novel insights into its mechanism of action.

## INTRODUCTION

Prostate cancer (PCa) is among the most common cancers in the United States and globally, with 313,780 new cases and 35,770 deaths in 2025 in the US alone (NCI SEER Cancer Stat Facts: Prostate Cancer, https://seer.cancer.gov/statfacts/html/prost.html)^1^. Globally, the World Cancer Research Fund International reported 1.47 million new PCa cases and 397,000 deaths from PCa in 2022 (Prostate Cancer Statistics, *World Cancer Research Fund*, 2024). While the majority of PCa tumors are adenocarcinomas, which are treatable through Androgen deprivation therapy (ADT or medical castration) and 2^nd^ generation androgen receptor (AR) antagonists^2–4^, 10-20% of PCa cases develop castration resistance within 5 years of the initial diagnosis, with this progression typically occurring after 2-3 years of hormonal manipulation^5,6^. When CRPC becomes metastatic (mCRPC), it no longer remains localized in the prostate and spreads to other tissues and organs like bone, lymph nodes, bladder, liver, lung, and brain, thus transforming into the most lethal subtype of PCa^6,7^. Taxanes are the first-line chemotherapeutics for mCRPC, but many patients develop taxane resistance, limiting the efficacy of this treatment^8^. Further, the presence of cancer stem cells (CSCs), like side populations and CD133+ cells (acquisition of mesenchymal phenotype or epithelial to mesenchymal trans-differentiation/EMT) with self-renewal and differentiation capacities, is believed to significantly contribute to the development of drug resistance^9^.

Given the increase in incidence of mCRPC and the growing issues with taxane resistance, novel drugs are needed to combat these trends. Drug re-purposing or repositioning to develop novel anti-cancer therapies has several advantages over drug discovery, including availability of extensive pre-existing knowledge of the drug’s safety, pharmacology, and toxicology, formulation, ease of manufacturing, and having already undergone several phases of preclinical and clinical studies for regulatory approval^10^. Furthermore, from a cost and time standpoint, re-purposing drugs that already have FDA approval for other disease states presents an important next step in chemotherapeutic development^11^.

Clofazimine (CLF) is a derivative of riminophenazines, FDA-approved for the treatment of leprosy, and is being repositioned for the treatment of multidrug-resistant tuberculosis^12^. We had previously identified CLF through high-throughput screening as a potentially efficacious agent for treating chronic myeloid leukemia (CML)^13^. Additionally, in a recently published study, we showed that CLF exhibits synergistic effect in combination with standard-of-care drugs in the treatment of multiple myeloma, specifically in PI and IMiD-resistant myeloma^14^. Interestingly, a phenotype-based screen identified CLF among several therapeutic agents selective for metastatic Prostate Cancer^13^. However, no other study so far has explored the functional consequences of CLF treatment specific to prostate cancer.

Due to the effectiveness of CLF against the aggressive, drug-resistant, and ‘stem-like’ forms of various cancers, this study focuses on the effects of CLF against a panel of PCa cell lines representing the diversity of patients with lethal and aggressive PCa. Furthermore, we show that CLF treatment downregulates several non-coding RNAs that have been shown to be associated with tumor cell proliferation, cell migration, invasion, and drug resistance in prostate cancer.

## MATERIALS AND METHODS

### Human Prostate Cancer Cell Lines

AR^lo^ mCRPC/NEPC cell lines (PC3, PC3M, DU145) and the osteotropic subline, C42b, were obtained from the American Type Culture Collection (ATCC; Manassas, VA, USA). The taxane-resistant cell line DUTXR was generated using dose escalation of taxanes over time. The cell lines were authenticated at the source and tested randomly at regular intervals for tissue specimen provenance and cell lineage at the AU Center for Pharmacogenomics and Single-Cell Omics (AUPharmGx) using the GenePrint 24 System (Promega; Madison, WI, USA). All cell lines are mycoplasma negative. The cell lines were maintained in an incubator at 37°C with 5% CO_2_ using the recommended ATCC media for each cell line supplemented with 10% FBS and Penicillin-Streptomycin.

### Drugs and Reagents

The drugs, reagents, and kits that were used are listed in **Table S1**. All drugs were solubilized in dimethyl sulfoxide (DMSO, Sigma-Aldrich; St. Louis, MO, US) and stored at −20°C.

### *In Vitro* Cytotoxicity Assays

PCa cell lines were treated with serial dilutions containing decreasing concentrations of CLF and DTX, as single-agent and combination therapy, for 48 hours. Cytotoxicity assays were performed using Resazurin cell viability assay (Ward’s Science; Rochester, NY, USA). Absorbance was measured using a Neo2 Microplate Reader (Biotek; Winooski, VT, USA). The half-maximal inhibitory concentration (IC_50_) values were calculated by nonlinear regression using a sigmoidal dose-response equation. These values were used to compare the dose response of single-agent and combination therapies. Drug synergy was calculated by comparing single-agent and combination drug-response data based on Chou-Talalay’s combination index (CI) method and the isobologram algorithm (CompuSyn software; Biosoft, US^15,16^. CI values between 0.9-0.3 and 0.3-0.1 signify synergism and strong synergism, respectively, between the drugs treated in combination.

### Apoptosis Assays

PCa cells were cultured for 48h in the presence of indicated concentrations of CLF and DTX as single agents and, in combination, harvested, washed, and incubated with Annexin V-FITC and Propidium Iodide for 15min at room temperature in the dark following the manufacturer’s protocol (BD Biosciences; Franklin Lakes, NJ, USA). Apoptosis was measured by BD LSR II flow cytometry (BD Biosciences; Franklin Lakes, NJ, USA) and analyzed using FlowJo™ Software (Ashland, OR, USA).

Caspase-3/7 activity assay was performed on the PCa cell lines using Caspase-Glo 3/7 luminescent assay kit according to the manufacturer’s instructions (Promega; Madison, WI, USA). Briefly, 1×10^6^/mL cells were cultured in 6-well plates and treated with either CLF alone or in combination with DTX for indicated times. After harvesting, cells were washed twice with cold PBS and resuspended with Caspase-glo 3/7 agents. Reactions were incubated for 1h at 37°C, and luminescence was determined using a Synergy 2 Microplate Reader (Biotek; Winooski, VT, USA). Cell death by apoptosis was also measured using immunoblotting analysis.

### Cellular Morphology Assessment

To assess <u>cellular morphology</u>, PCa cells were seeded at 0.025*10^6^ cells/ml in 6-well plates and exposed to single-agent DTX, CLF, or DTX+CLF combination for 48 hr. Three areas with approximately equal cell densities were identified in each well, and images were captured with an EVOS FL digital cell imaging system (Thermo Fisher Scientific, Inc.) using a 10X□objective.

### Cell migration (Scratch) Assay

Cells were plated in 6-well plates at 1×10^5^ cells/well and incubated for 48hr to a 95% confluency. The monolayer was scratched with a SPLScar Scratcher 6-well tip at a width of 0.50 mm at the center of the well. CLF or DTX single agents or DTX+CLF combination doses were applied to the cells. PCa cell culture medium supplemented with 10% FBS containing 0.05 % DMSO was added to the cells in the control wells (vehicle). Micrographs of the wound areas were obtained at 0, 24, and 48hr using an EVOS FL digital cell imaging system (Thermo Fisher Scientific, Inc.). The images were captured in brightfield and phase contrast modes at 20X and 40X magnifications. The area of the initial wound (at 0 hr) and the “gap area” were measured at 48hr with Image J software.

### Microfluidic Cell Migration Assay

The fabrication of a Polydimethylsiloxane (PDMS)-based microchannel assay using standard multilayer photolithography and replica molding has been demonstrated earlier^17,18^. For this study, PCa cells were seeded in 6-well plates and treated with 10 μM and 25 μM CLF. Following 24-hour incubation, 2×10^6^ cells/mL were introduced into the cell seeding inlet line of the microfluidic channel via pressure-driven flow and were allowed to adhere for 30-40 min at 37 ^°^C, 5% CO_2_. Next, a chemotactic gradient was established to induce cell entry into the microchannels by adding serum-free media to the cell seeding inlet and media supplemented with 10% FBS to the chemoattractant inlet. The devices were placed on an automated Nikon Ti2 Inverted Microscope equipped with a Tokai Stage-Top incubator unit, which maintained cells at 37 °C and 5% CO_2_. Time-lapse Images were captured every 10 minutes for 8 hours with a 10x /0.45 NA Ph1 objective. The percentage of cell entry into the microchannels was calculated by dividing the number of cells that entered the microchannels by the total number of cells located within 50□μm of the microchannel entrance.

### Aldeflour Activity Assay

Aldehyde dehydrogenase (ALDH) activity was assessed using the ALDEFLUOR ™ assay kit according to the manufacturer’s instructions (Stem Cell Technologies; Vancouver, Canada). Briefly, 1×10^6^ PCa cells were treated with CLF-based regimens for 24h, harvested, and resuspended in 1 mL Aldefluor assay buffer containing the ALDH substrate BODIPY-aminoacetaldehyde (BAAA). Negative control samples were treated with 5 µL of diethylaminobenzaldehyde (DEAB) as an inhibitor of ALDH1 enzymatic activity. Cells were incubated for 30-45 minutes at 37 °C, washed twice, and suspended in Aldefluor assay buffer. The brightly fluorescent ALDH+ cells were detected by BD LSR II flow cytometer (BD Biosciences; Franklin Lakes, NJ, USA) and analyzed using FlowJo™ Software (Ashland, OR, USA).

### Detection of the Mitochondrial Membrane Potential (MMP)

Untreated, DTX-treated, CLF-treated, and DTX+CLF combination-treated (24hrs) cells were incubated with 5μM JC-1 dye for 15 min in the dark at 37°C and washed twice in PBS. Mitochondrial Membrane Potential was then assayed using Synergy Neo2 Microplate Reader (Biotek).

### DCFDA Total Cellular Reactive Oxygen Species (ROS) assay

PCa cells were treated for 24hrs with CLF. Following 24hrs, cells were incubated with 10μM DCFDA in RPMI (Phenol red-free) medium at 37°C for 30min, washed twice with phosphate buffer saline (PBS). ROS production was measured using Synergy Neo2 Microplate Reader (Biotek).

### Gene Expression Profiling (GEP) Analysis

Cells were plated at a density of 4×10^5^/ml cells and were treated with CLF. After 24h incubation, high-quality RNA was isolated using QIAshredder and RNAeasy kit (Qiagen; Hilden, Germany). RNA concentration and integrity were determined using the Nanodrop-8000 (Thermo-Fisher Scientific; Waltham, MA, US) and Agilent 2100 Bioanalyzer (Agilent Technologies; Santa Clara, CA, US) and stored at −80 °C. An RNA integrity number (RIN) threshold of 8 was used for RNA-seq analysis. RNA-seq libraries were constructed using Illumina TruSeq RNA sample preparation kit v2 (Illumina; San Diego, CA). Next-generation RNA sequencing was performed on Illumina’s NovaSeq platform using a 150bp paired-end protocol with a depth of > 20 million reads per sample.

### RNAseq Data Analysis

Gene expression profiling (GEP) data was pre-processed and filtered (genes with mean counts<10 were removed), and the normalized (CPM - counts per million) data was analyzed using the Partek Flow package (Partek, Inc.; St. Louis, MO, USA) to perform differential expression testing to identify GEP signatures. Least squares (LS) mean values were calculated for each group as the linear combination or sum of the estimated means from the linear model. LS mean is model-dependent and produces a more accurate, unbiased estimate of the group means even in unbalanced data. An LS Mean threshold of >=1 was applied for analysis, and differential gene expression analysis between groups was performed using limma, an empirical Bayesian method, to detect differentially expressed (DE) genes. The advantage of limma compared to traditional t-tests is that limma provides moderated t-test statistics by shrinking the variance statistics, therefore improving the statistical power^14^. Mean fold-change>|1| and p<0.05 were considered as the threshold for reporting significant differential gene expression. Heatmaps were generated using unsupervised hierarchical clustering (HC) analysis based on the top DE genes.

### Pathway analysis

Based on the most significant DE genes, Ingenuity Pathway Analysis (IPA) software (Qiagen; Hilden, Germany) was used to identify i) the most significantly affected molecular pathways, ii) upstream regulator molecules like microRNA and transcription factors, iii) downstream effects and biological processes, and iv) causal networks predicted to be activated or inhibited in response to treatment^19^.

Further, pathway analysis was also performed using Gene Set Enrichment Analysis (GSEA) and Gene Set ANOVA, a statistical method that uses analysis of variance (ANOVA) to detect changes in functional groups of genes (pathways) based on the list of differentially expressed genes against the MSigDB database enrichment background.

Finally, we used Pathway Figure OCR, an open science tool from the Wikipathways platform for biological pathways, to extract pathway information from published literature and performed enrichment analysis using WebGestalt to demonstrate the relevance of the top gene sets^20^.

### Single-cell RNA sequencing (scRNAseq)

Automated single-cell capture and cDNA synthesis were performed on the mCRPC cell lines (n=4: PC3, PC3M, DU145, and DUTXR) using the 10X Genomics Chromium Controller with the Chromium Next GEM Single Cell 5’ Gene Expression (Dual Index) kit. Single-cell RNA sequencing was performed on Illumina HiSeq 2500 NGS platform (Paired-end, 2*125bp, 100 cycles, v3 chemistry) at ∼50,000 paired-end reads per cell.

### scRNAseq data analysis

scRNAseq datasets were obtained as matrices in the Hierarchical Data Format (HDF5/H5). Demultiplexing of sequencing results, barcode processing, read alignment, and UMI counting were performed using the 10X Cell Ranger analysis pipeline v3.1. Reads were aligned to the genome reference GRCh38, and Partek Flow and Seurat R packages were used to perform data QC, feature selection, dimension reduction, unsupervised clustering, and differential expression analyses (DEA). Cells with <100 genes detected or >40% of mitochondrially-encoded transcript counts and genes detected in <20% of cells were excluded before analysis. Principal components analysis (PCA) was performed on the integration-transformed expression matrix, and the first 100 principal components (PC) were used for Leiden clustering of cells with a resolution parameter of 1. Highly variable genes were selected for clustering analysis based on a graph-based cluster approach. Visualization of cell populations was performed by T-distributed stochastic neighbor embedding (t-SNE) on the same PCs with 100 nearest neighbors. Expression patterns associated with subpopulations were identified using single-cell-level marker gene expression^21^.

### Patient Datasets

Gene expression data on Prostate Adenocarcinoma patients included in The Cancer Genome Atlas (TCGA) database (TCGA-PRAD) was extracted from the Genomic Data Commons (GDCs) server (cancergenome.nih.gov). The interactive web-based tool Gene Expression Profiling Interactive Analysis (GEPIA) was used for in-depth analysis of TCGA gene expression data files and to compare transcriptome data on target candidate pathway genes with patient survival (overall survival (OS) or disease-free survival (DFS)) from the prostate expression data matrix^22^. Further, raw RNAseq data on PCa patients were downloaded from the Gene Expression Omnibus database (GSE54460; BioProject ID PRJNA237581)^23^. This dataset consists of formalin-fixed paraffin-embedded (FFPE) prostatectomy samples from 100 PCa patients (49 with BCR, 51 with no BCR) from 3 independent sites: the Atlanta VA Medical Center, Moffitt Cancer Center, and Sunnybrook Health Sciences Centre at the University of Toronto.

### Statistical Analysis

All statistical analysis was performed using R v4.3.2 and GraphPad Prism v10. All tests were two-sided, with p<0.05 being considered statistically significant.

## RESULTS

### CLF treatment results in loss of viability and increased apoptosis in PCa cell lines

The effectiveness of CLF was first evaluated using *in vitro* cytotoxicity assays in our mCRPC cell line panel (AR mCRPC; PC3, PC3M, DU145, taxane-resistant DUTXR, and AR^Hi^ mCRPC; C4-2B), which represents the broad spectrum of biological and genetic heterogeneity of metastatic PCa patients, as well as innate and acquired taxane resistance. First, using *in vitro* cytotoxicity assay, we confirmed that single-agent CLF treatment showed a reduction in cell viability following 48h of treatment (**Figure S1**), which was further enhanced when CLF was combined with DTX (**Figure 1A**). Drug synergy analysis using Chou-Talalay’s combination index (CI) method showed a CI value of <0.3, indicating strong synergism^15,16^. Furthermore, the Dose reduction index (DRI) for DTX was >20 for the docetaxel-resistant PCa line DUTXR, suggesting that combination therapy with CLF reduces the dose requirement of DTX by >20-folds. Next, we showed that CLF treatment reduced the cell density of mCRPC cell lines. Cells were exposed to DTX and CLF treatment as single agents, and the DTX+CLF combination and cellular morphology were assessed using phase-contrast microscopy. Concurrent with our *in vitro* cytotoxicity assay results, micrographs of PCa cells exposed to both CLF and CLF+DTX treatment showed a decrease in cell density compared to control cells, as shown in **Figure 1B**. Finally, we tested the impact of CLF treatment on cellular apoptosis using Annexin V-PI staining. We observed that treatment with CLF as a single agent resulted in elevated levels of apoptosis in PCa lines compared to no-drug control. Further, the percentage of apoptotic cells in each cell line tested was significantly higher following CLF+DTX combination treatment compared to single-agent treatment (**Figure 1C**). Our results were confirmed using Caspase 3/7 activity assay (**Figure 1D**).

**Figure 1.**
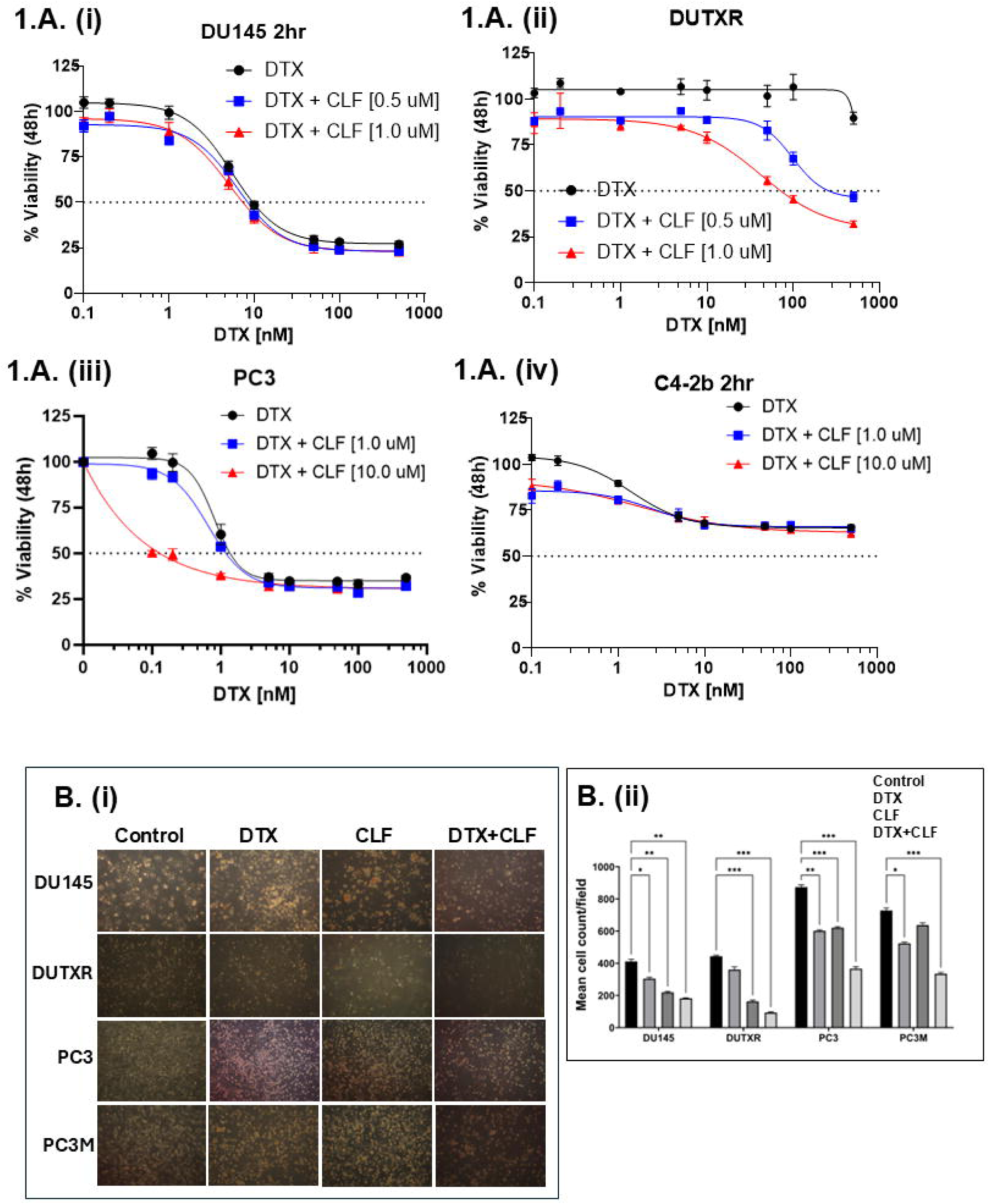
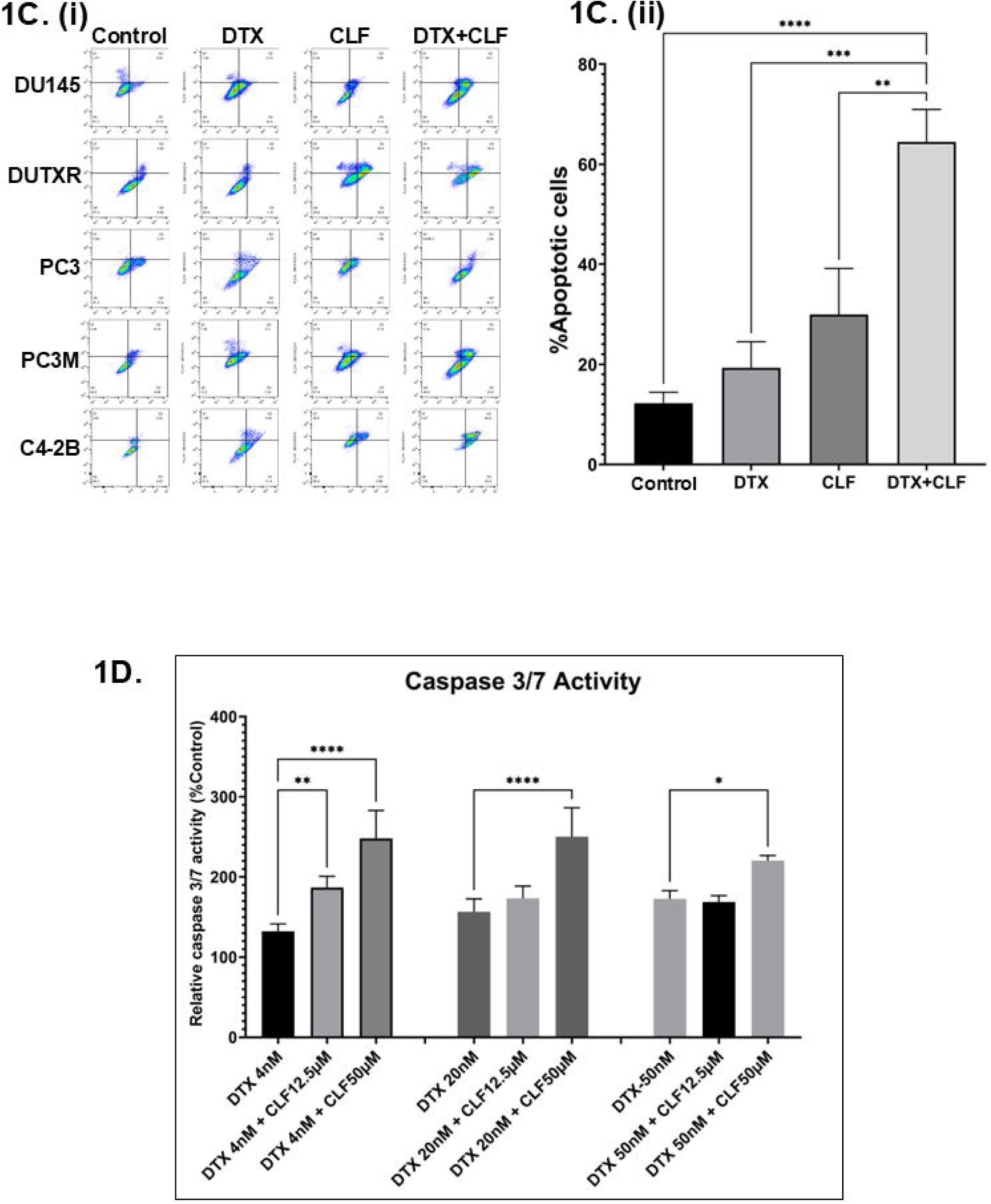
*In vitro* cell viability assays. A. Representative dose-response plots showing *in vitro* cytotoxicity of Clofazimine in combination with Docetaxel (DTX) in the mCRPC cell line pair DU145 and DU-TXR (i-ii), PC3 (iii), and C4-2b (iv). Chemosensitivity assays were performed using Resazurin Assay. **B. Assessment of cellular morphology and cell density:** (i) Representative images showing the effect of DTX and CLF drugs on cell count and cell morphology of mCRPC cells. The images of cells were captured before (0h) and after (48h) CLF treatment either as a single agent or in combination. (ii) Microscopy results show significantly higher cell death in combination treatment compared to single-drug treatment for all cell lines; ImageJ data analysis showed a significant difference in cell density for both CLF single-agent and DTX+CLF combination treatments. Results show significantly higher cell death in combination treatment compared to single-drug treatment for mCRPC cell lines (Significant value * = p ≤ 0.05). **C. % Apoptotic cells representing the effect of CLF on PCa cell lines as a single agent and in combination with DTX.** Cells were treated for 48hr, and then apoptosis was measured using Annexin V-FITC and flow cytometry. **D. Representative Caspase 3/7 activity assay plots showing increased apoptosis following CLF treatment.**

### Cell migration/Scratch Assay

To find if CLF treatment induced reduction of mCRPC cell migration, we performed scratch assays by creating a “scratch” in the cell monolayer and treating it with a single dose of CLF, DTX, and CLF+DTX combination. Images were captured at the beginning (0 hr) and at regular intervals (24 and 48hr), and the migration rate of the cells to close the scratch was quantified. Our results showed that cell migration was higher in control cells compared to drug-treated cells (**Figure 2A**). Further, combination treatment (CLF+DTX) showed a higher impact on reducing cell migration in PCa cell lines compared to treatment with single agents. Scratch assay is also an indicator of the effect of CLF on the metastatic spread, resistance to therapy, as well as ‘cancer stemness’ potential of PCa cells.

**Figure 2.**
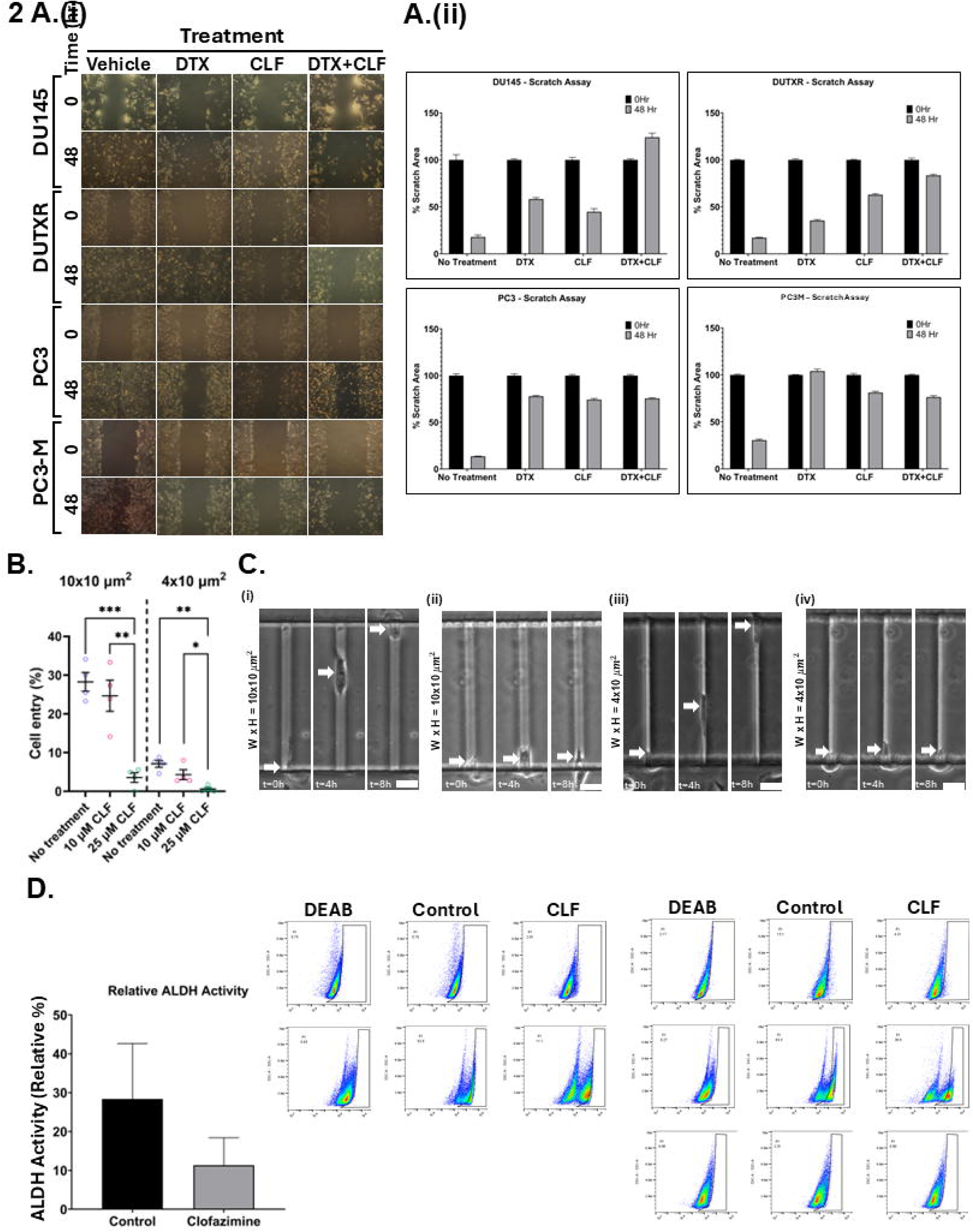
CLF is potentially effective against cancer metastasis and ‘stemness’ in lethal PCa. A. Wound healing/Scratch assay: (i) Representative plots showing results of wound healing (Scratch) assay. Cell migration after 24hr and 48hr CLF single-agent and CLF+DTX combination were assessed by measuring the scratch size. Images were captured before (0h) and after (24hr and 48hr) drug treatments (Significance P-value * = p ≤ 0.05). (ii) Bar graphs showed a significant reduction in cell migration (wound healing) following CLF-based single-agent and combination treatments. **B. Cell entry and migration in microfluidic devices.** The fabrication of a μ-channel assay is described in the *Methods* section. Reduced cell entry into channels indicates a more generic defect in the capacity of tumor cells to invade physically restricted spaces.. (i) Percentage of DU-145 cells, untreated or treated with 10 μM or 25 µM CLF, that enter moderately (*W(idth)* x *H(eight)* = 10 x 10 μm^2^ and laterally (*W* x *H* = 4 x 10 μm^2^) confined microchannels (4 independent experiments). **C. Representative images of control and CLF-treated prostate cancer cells migrating through** μ**-channels:** (i, iii) Image sequence showing untreated DU-145 cells migrating through (i) moderately or (iii) laterally confined microchannels. (ii, iv) Image sequence showing that treatment with 25 μM CLF prevents DU-145 cells from entering (ii) moderately or (iv) laterally confined microchannels. Statistical tests: (i) one-way ANOVA followed by Tukey’s multiple-comparisons post hoc test. \**p*<0.05, \*\**p*<0.01, \*\*\**p*<0.001 Values represent mean ± SEM (i). Scale bar: 20 µm. **D. Aldefluor (ALDH) activity in cell lines following CLF treatment.** i) Cells were treated for 24 hr, and ALDH activity was measured using the Aldefluor assay kit and flow cytometry (DEAB: positive control). The brightly fluorescent ALDH+ cells were detected by BD LSR II flow cytometry. ii) Barplots representing mean relative ALDH activity compared to DEAB (negative control) across PCa lines.

### CLF reduces the migration of mCRPC cells through microfluidic channels

The Polydimethylsiloxane-PDMS-based microchannel assay served as a physiologically relevant *in vitro* metastasis model that allowed us to study the effect of CLF on tumor cell motility through microchannels of dimensions that mimic the size of the tracks encountered by migrating cells *in vivo*^24,25^. To assess the migration efficiency of CLF-treated mCRPC cells compared to controls, we calculated the percentage of cell entry into the microfluidic channels, defined as the total number of cells entering the channels divided by the total number of cells seeded within a distance of 50 μm from the microchannel entrances. We observed that CLF treatment nearly abolished cell entry of PCa cells into both moderately (*W(idth)*x*H(eight)* = 10×10μm^2^) and laterally (*W*x*H* = 4×10μm^2^) confined microchannels, indicating its potential to suppress PCa cell invasion and metastasis (**Figure 2B** and **Videos**).

### CLF inhibited aldehyde dehydrogenase activity in PCa cells

ALDH has been shown to be associated with ‘cancer stemness’ and resistance to taxane therapy in prostate cancer. **Figure 2C** shows higher baseline aldehyde dehydrogenase (ALDH) activity level in the clonally derived taxane-resistant mCRPC cell line DUTXR compared to the parental line (DU145). This is a significant finding as ALDH is an intracellular enzyme that is frequently over-expressed in cancer stem cells and has been linked to drug resistance. Further, we showed (**Figure 2C**) that treatment with CLF reduced ALDH activity in the PCa cell lines tested when compared to the no-treatment control.

### Gene expression profiling revealed coding and non-coding genes associated with CLF treatment in lethal PCa

Next, we performed genome-wide transcriptome (next-generation mRNA sequencing or RNAseq) analysis to identify genes that were differentially regulated following CLF treatment. Volcano plots showing these differentially expressed (DE) genes are provided in **Figure 3A**. Among these, 557 genes were shown to be significantly differentially regulated following CLF treatment (p<0.05; fold-difference ≠ 1; 260 up-regulated and 297 down-regulated). Of these genes, 497 had an absolute fold-change ≥ 2 (243 Up-regulated and 254 down-regulated). The Venn diagram in **Figure 3B** shows that among the genes that were differentially regulated following CLF treatment (fold change ≠ 1), 1027 were common between all the mCRPC cell lines (PC3, PC3M, and DU145). **Figure 3C** shows a heatmap representing the top genes that were significantly differentially expressed between CLF-treated vs untreated controls. **Supplementary Table S2** lists the top coding genes with log_2_ratio (CLF vs treatment) >|2|. The top differentially expressed genes in the DUTXR cell line are listed in **Supplementary Table S3**.

**Figure 3.**
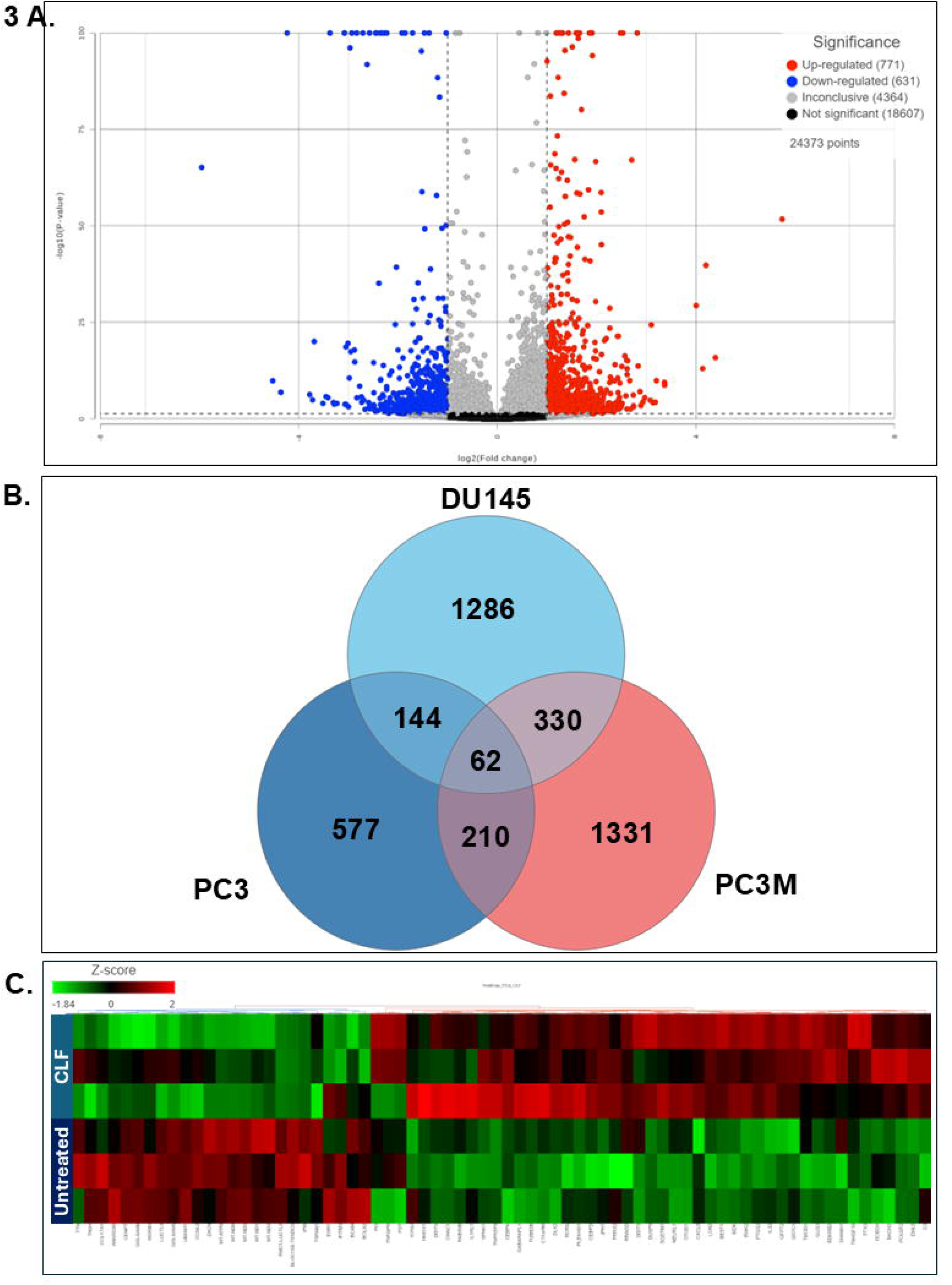
Gene expression analysis: Control vs CLF treatment. **3A.** Volcano plot showing the differentially expressed (DE) genes. **3B.** Venn Diagram showing CLF-regulated genes that were common between the cell lines. **3C**. Heatmap representing the top genes that were most significantly differentially expressed between CLF-treated vs untreated controls.

### Pathway analysis revealed top differentially regulated molecular pathways associated with PCa cells treated with CLF

A graphical summary of the IPA analysis based on differentially regulated genes following CLF treatment in PCa cell lines is shown in **Figure 4A**, where orange indicates activation and blue indicates inhibition. Top differentially regulated canonical pathways associated with CLF treatment include oxidative phosphorylation, autophagy, rRNA processing, mitochondrial dysfunction, cancer cell movement/metastasis, inflammatory response, and cellular homeostasis (**Figure 4B**). Additional pathway analyses using GSEA and Gene set ANOVA against the background of KEGG and mSigDB databases confirmed the downregulation of oxidative phosphorylation and upregulation of autophagy as top CLF treatment-induced pathways (**Figure 4C**). Further, WebGestalt analysis using the Reactome database showed downregulation of interferon alpha/beta signaling as another top-downregulated pathway (Normalized Enrichment Score 2.677; FDR<0.001) while the top WikiPathways Cancer pathway was downregulation of Epithelial to mesenchymal transition (EMT), which is a hallmark of ‘stemness’ ^26^. **Figure 4D** represents the top upstream regulators predicted by IPA to be activated or inhibited following CLF treatment. These include ATF4, TWNK, and LONP1. ATF4, a member of the ATF/cAMP response element-binding (CREB) family, is known to play key roles in cellular response to stress, particularly endoplasmic reticulum (ER) stress and oxidative stress, promotes autophagy, and is involved in cell cycle arrest and apoptosis ^27,28^, while lon peptidase 1 (LONP1) and the mitochondrial helicase TWNK are involved in controlling mitochondrial function ^29,30^.

**Figure 4.**
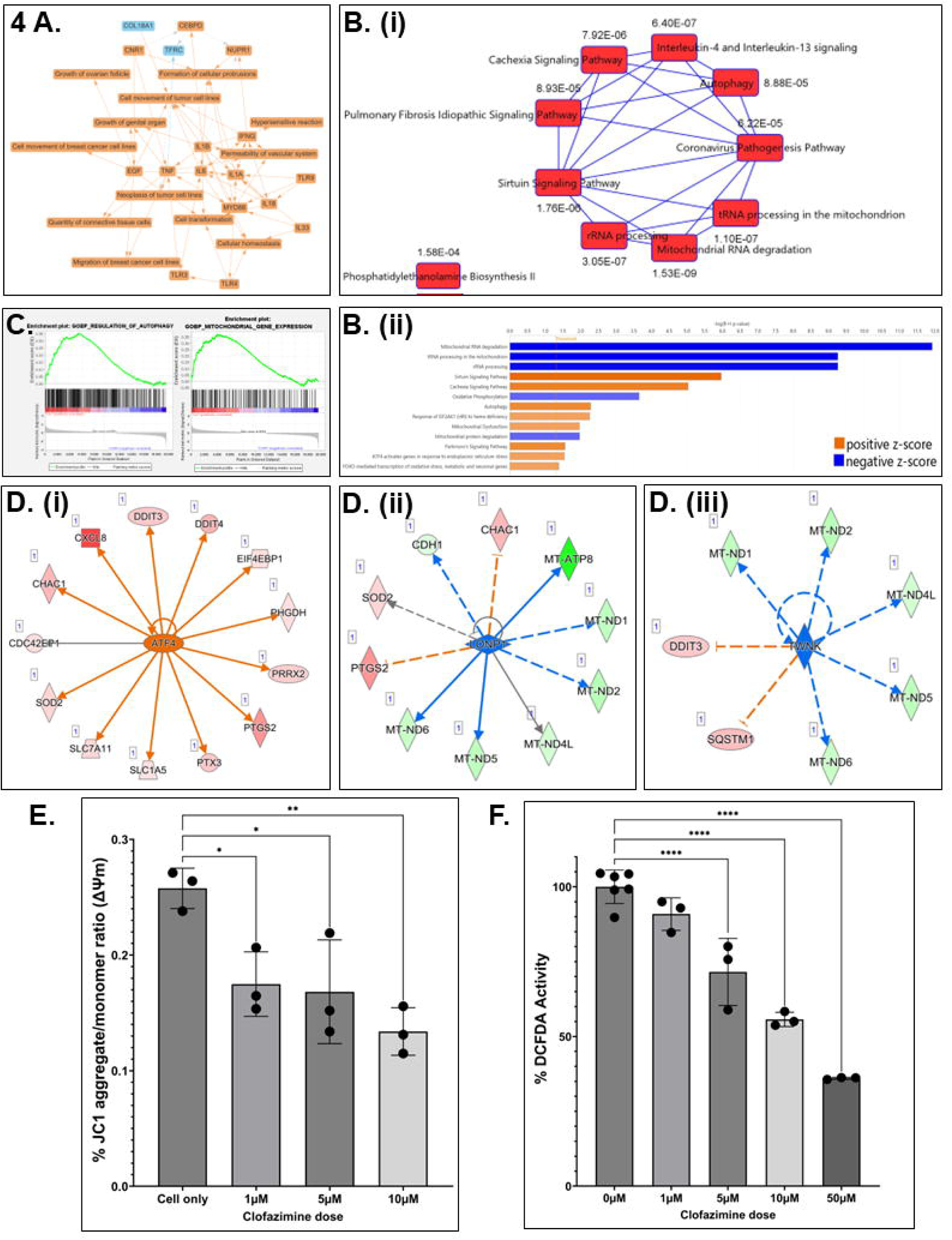
Pathway analysis based on differentially expressed genes. **4A**. Graphical Summary of Ingenuity Pathway Analysis (IPA). **B. (i-ii)** Top Canonical Pathways predicted using IPA (B-H>1.3; |z|>2) and **C**. GSEA results showing top dysregulated pathways. **D**. (i-iii)Top predicted Upstream regulators (p-value of overlap <0.05; Predicted activation z-score & expression fold change>|1.5|). **E-F. CLF affects mitochondrial membrane potential (MMP) and intra-cellular reactive oxygen species (ROS) generation in PCa cells**. Representative figures showing the DU145 cell line. **E. Mitochondrial membrane potential (MMP):** JC-1 (Sigma), a cationic carbocyanine dye that accumulates in mitochondria, was used to assess Mitochondrial membrane potential (ΔΨ_M_). The JC-1 aggregate (healthy mitochondria) vs monomer (depolarized mitochondria) ratio is a measure of (ΔΨ_M_)**. F. Reactive Oxygen Species (ROS) generation:** DCFDA (Sigma) was used to measure intracellular ROS production by flow cytometry. DCFDA shows fluorescence when oxidized.

### CLF treatment results in altered mitochondrial metabolism

Next, to confirm the findings of pathway analysis, we analyzed CLF treatment-induced changes in mitochondrial membrane potential using JC-1 (Sigma). JC-1 is a cationic fluorescent dye that accumulates at a higher level in the mitochondria of cancer cells, representing elevated mitochondrial membrane potential (ΔΨ), which may be the result of enhanced oxidative phosphorylation. This accumulation of JC-1 leads to the formation of J-aggregates, which can be assayed through red fluorescence (emission maximum at 590 nm) compared to the green fluorescence of J-monomers. Further, we used DCFDA (Sigma) to measure intracellular ROS levels and confirm the effect of CLF on reactive oxygen species (ROS), such as superoxides and hydrogen peroxide (H_2_O_2_). Significantly, we observed that CLF treatment results in a decrease of JC1 red/green ratio, indicating mitochondrial membrane dysfunction and depolarization (**Figure 4E**). We also showed that CLF treatment resulted in a decrease of DCFDA signal, indicating a reduction of intracellular ROS production (**Figure 4F**). This antioxidant effect of CLF is unique to prostate cancer, since we have earlier observed an increase in ROS production following CLF administration in multiple myeloma and CML cells^14,31^.

### Single-cell transcriptomics (scRNAseq) showed high expression of top non-coding RNAs MALAT1 and NEAT1 in mCRPC cells

Figure 5A.**i** displays t-SNE clusters generated from baseline (untreated) scRNAseq data on individual single cells in mCRPC cell lines (∼3500 cells). Each dot represents a single cell. We have earlier demosttrated that AR^low^ cells (primarily belonging to the mCRPC subtype) show higher expression of several mesenchymal gene signatures involved in Epithelial-mesenchymal transition (EMT), including Vimentin, N-cadherin (CDH2), Fibronectin (FN1), S100A4, Snail (SNAI1), Slug (SNAI2), CDH11, TWIST1, and ZEB1, as well as the upregulation of cancer stemness-related markers like Urokinase-type plasminogen activator (PLAU), Urokinase-type plasminogen activator receptor (PLAUR), and CD44, which were further up-regulated in the taxane-sensitive mCRPC cell line DU145-TXR, in addition to the drug resistance markers CDK1 and CXCL8^32^. Here, through investigation of the transcriptome profiles of non-coding RNAs in individual cells within mCRPC cell lines, we observed a significantly large proportion (>90%) of subclones with high expression (log_2_(CPM)>1) of the top non-coding RNAs MALAT1 and NEAT1 among all 4 cell lines (Figure 5A**.ii-5B**).

**Figure 5.**
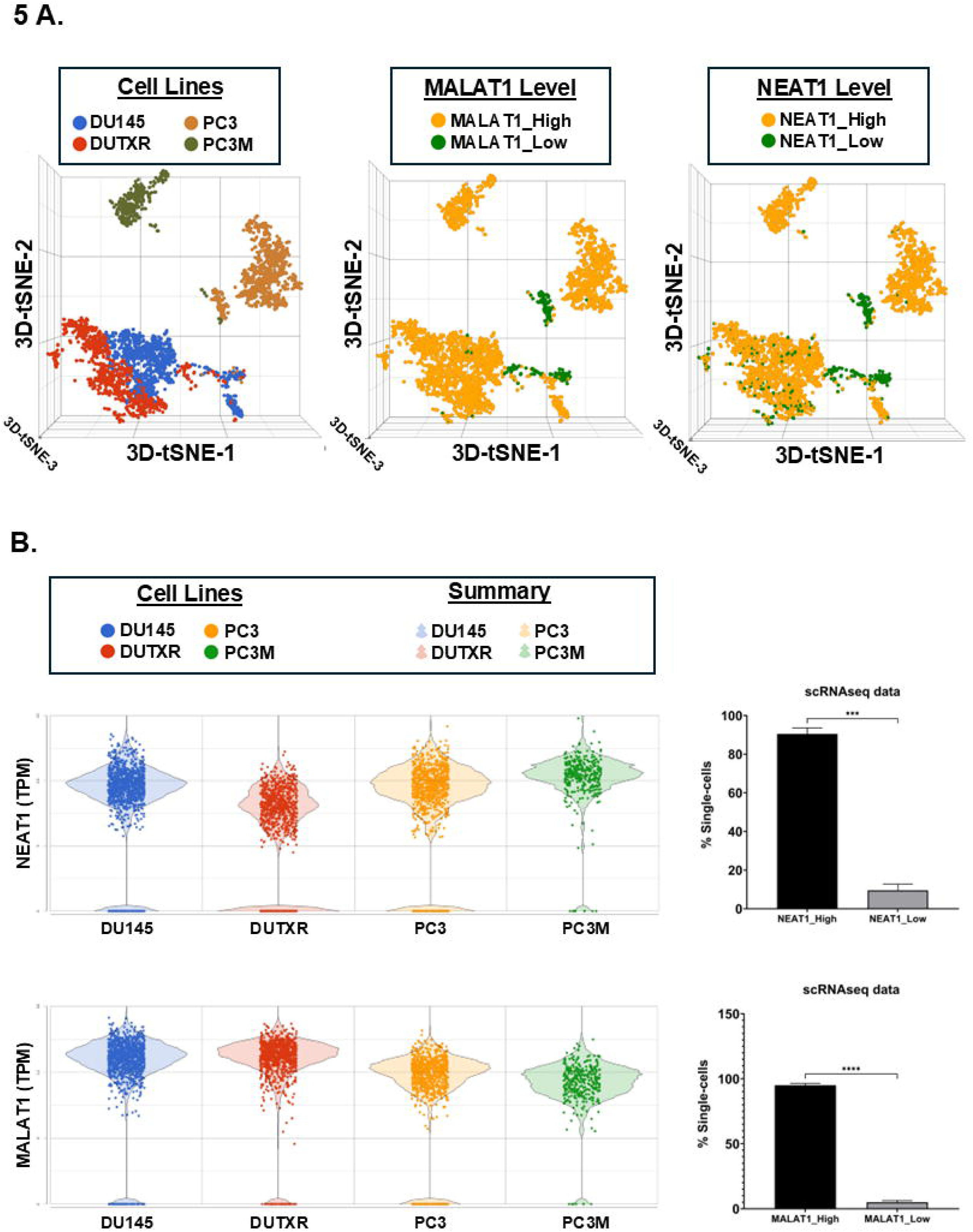
Single-cell transcriptomics identifies enrichment of non-coding RNA signatures in metastatic prostate cancer cells. A. t-distributed stochastic neighbor embedding (t-SNE) plots and B. Bar plots showing the comparison between the single-cell clusters representing the mCRPC cell lines (n=4; PC3, PC3M, DU145, DUTXR). We observed higher proportion of single cells showing enrichment of the expression of non-coding RNAs. Single-cell capture and scRNAseq data analysis are described in the *Methods* section. Single-cell RNA sequencing was performed using the 10X Genomics Chromium Platform. Each dot represents a single cell. Contaminated (doublet) cells were not included.

### CLF treatment-induced differential regulation of Non-coding RNAs was validated using patient datasets

Notably, non-coding genes, including lncRNAs and snRNAs, that were significantly associated with CLF treatment in mCRPC cell lines are listed in **Table 1**. The top differentially regulated non-coding RNAs that are most relevant to this study are represented using a heatmap (Figure 6A) and bar plots (Figure 6B).

**Figure 6.**
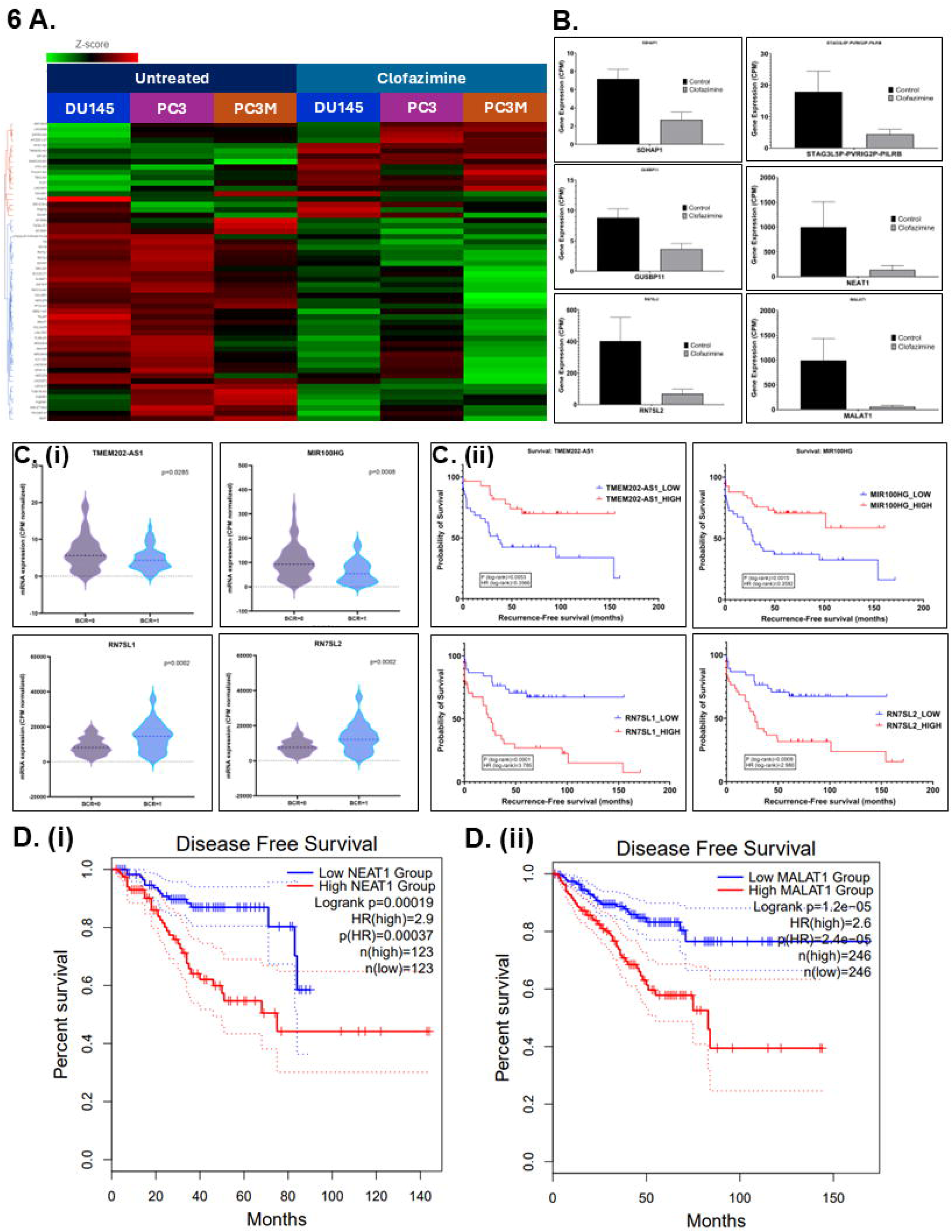
Top lncRNAs and snRNAs are differentially regulated following CLF treatment in mCRPC cell lines. **A.** Heatmap (Details are provided in **Table 1**); **B.** Box plots showing the differential expression of top non-coding RNAs**; C-D. Validation of CLF treatment-related non-coding RNA signatures using patient datasets**. **C. Clinical data (GSE54460)** (i) Violin Plots and (ii) Kaplan-Meier Curves show that the top CLF treatment-induced non-coding genes are significantly associated with Biochemical Recurrence (BCR) in PCa patients, respectively. **D. TCGA’s prostate adenocarcinoma (PRAD) GEP dataset.** Kaplan-Meier Curves show that the top CLF treatment-induced non-coding RNAs (i) NEAT1 and (ii) MALAT1 are significantly associated with disease-free survival in PCa patients.

**Table 1:** Top differentially regulated non-coding RNAs following CLF treatment (p<0.05).

| Gene name | Gene ID | Gene biotype | P-value (CLF vs No treatment) | Fold change (CLF vs No treatment) |
| --- | --- | --- | --- | --- |
| **MALAT1** | ENSG00000251562 | lncRNA | 1.07E-170 | -18.84 |
| **RN7SK** | ENSG00000283293 | snRNA | 1.30E-05 | -13.20 |
| **NEAT1** | ENSG00000245532 | lncRNA | 4.06E-159 | -10.36 |
| **TUBA1B-AS1** | ENSG00000258017 | lncRNA | 1.88E-04 | -8.32 |
| **LINC00342** | ENSG00000232931 | lncRNA | 2.69E-11 | -7.91 |
| **RN7SL1** | ENSG00000276168 | misc_RNA | 6.84E-97 | -7.83 |
| **MT-RNR1** | ENSG00000211459 | Mt_rRNA | 0.00E+00 | -6.54 |
| **RN7SL2** | ENSG00000274012 | misc_RNA | 1.40E-92 | -6.17 |
| **MBNL1-AS1** | ENSG00000229619 | lncRNA | 2.03E-04 | -4.86 |
| **TGFB2-OT1** | ENSG00000281453 | lncRNA | 1.93E-02 | -4.63 |
| **NSUN5P1** | ENSG00000291151 | lncRNA | 1.21E-06 | -4.12 |
| **WASH5P** | ENSG00000292982 | lncRNA | 2.18E-07 | -3.93 |
| **7SK** | ENSG00000202198 | misc_RNA | 1.32E-03 | -3.89 |
| **TALAM1** | ENSG00000289740 | lncRNA | 2.11E-02 | -3.76 |
| **HERC2P9** | ENSG00000291082 | lncRNA | 7.76E-04 | -3.48 |
| **FLNB-AS1** | ENSG00000244161 | lncRNA | 2.36E-02 | -3.35 |
| **FKBP9P1** | ENSG00000291029 | lncRNA | 5.83E-03 | -3.31 |
| **MIR23AHG** | ENSG00000267519 | lncRNA | 3.20E-07 | -3.27 |
| **STAG3L5P-PVRIG2P-PILRB** | ENSG00000272752 | lncRNA | 2.96E-04 | -3.17 |
| **MUC20-OT1** | ENSG00000242086 | lncRNA | 2.16E-05 | -3.13 |
| **MIR222HG** | ENSG00000270069 | lncRNA | 1.28E-02 | -3.12 |
| **PDCD6IP-DT** | ENSG00000271643 | lncRNA | 2.12E-03 | -3.03 |
| **RNF213-AS1** | ENSG00000263069 | lncRNA | 3.89E-02 | -3.03 |
| **GOLGA2P5** | ENSG00000290571 | lncRNA | 1.88E-02 | -2.89 |
| **RTCA-AS1** | ENSG00000224616 | lncRNA | 1.87E-02 | -2.89 |
| **KLC1-AS1** | ENSG00000246451 | lncRNA | 4.07E-02 | -2.77 |
| **USP34-DT** | ENSG00000270820 | lncRNA | 1.75E-02 | -2.70 |
| **MIRLET7BHG** | ENSG00000197182 | lncRNA | 4.02E-02 | -2.69 |
| **MT-RNR2** | ENSG00000210082 | Mt_rRNA | 0.00E+00 | -2.60 |
| **ZNF767P** | ENSG00000291118 | lncRNA | 1.24E-02 | -2.58 |
| **ERVK13-1** | ENSG00000260565 | lncRNA | 9.18E-03 | -2.34 |
| **SDHAP1** | ENSG00000290763 | lncRNA | 2.81E-02 | -2.28 |
| **LINC-PINT** | ENSG00000231721 | lncRNA | 9.68E-04 | -2.28 |
| **GUSBP11** | ENSG00000228315 | lncRNA | 2.25E-02 | -2.17 |
| **WAC-AS1** | ENSG00000254635 | lncRNA | 7.46E-05 | -2.06 |
| **LINC03072** | ENSG00000273270 | lncRNA | 3.62E-02 | -2.05 |
| **TRIM16L** | ENSG00000291110 | lncRNA | 4.08E-03 | 1.88 |
| **MIR100HG** | ENSG00000255248 | lncRNA | 4.34E-03 | 1.95 |
| **KTN1-AS1** | ENSG00000186615 | lncRNA | 1.68E-03 | 2.02 |
| **LINC00886** | ENSG00000240875 | lncRNA | 2.20E-02 | 2.22 |
| **MIR1915HG** | ENSG00000204682 | lncRNA | 3.40E-02 | 2.26 |
| **ZNF503-AS2** | ENSG00000237149 | lncRNA | 1.00E-03 | 2.44 |
| **TBX2-AS1** | ENSG00000267280 | lncRNA | 4.39E-02 | 2.75 |
| **SMARCA5-AS1** | ENSG00000245112 | lncRNA | 4.45E-04 | 2.93 |
| **APCDD1L-DT** | ENSG00000231290 | lncRNA | 7.17E-03 | 2.98 |
| **MYG1-AS1** | ENSG00000257605 | lncRNA | 2.26E-02 | 3.00 |
| **PCAT1** | ENSG00000253438 | lncRNA | 7.60E-05 | 3.84 |
| **LINC00973** | ENSG00000240476 | lncRNA | 9.36E-07 | 3.92 |
| **TMEM202-AS1** | ENSG00000261423 | lncRNA | 7.54E-05 | 4.79 |
| **PHLDA1-AS1** | ENSG00000257453 | lncRNA | 4.28E-05 | 7.63 |
| **MIF-AS1** | ENSG00000218537 | lncRNA | 3.38E-06 | 7.83 |

RNAseq data on PCa patients were obtained from the Gene Expression Omnibus database (GSE54460; BioProject ID PRJNA237581)^23^. This dataset includes 100 PCa patients (49 with biochemical recurrence (BCR), 51 with no BCR) from the Atlanta VA Medical Center, Moffitt Cancer Center, and Sunnybrook Health Science Center. We performed differential gene expression analysis and observed that the expression of several top non-coding RNAs was associated with survival and biochemical recurrence (BCR) in these patients (Figure 6C).

Further *in silico* analysis of the non-coding gene signature using TCGA’s prostate adenocarcinoma (PRAD) GEP dataset showed that several non-coding RNAs that were significantly differentially dysregulated following CLF treatment were also significantly associated with disease-free survival (Kaplan-Meier curves; Figure 6D), with Hazards Ratios >2 and p<0.05. Thus, these biomarkers may serve as additional cues to the mechanism of action of Clofazimine, particularly its efficacy in metastatic/aggressive PCa.

## DISCUSSION

Drug resistance severely limits the efficacy of current chemotherapeutics, with the exact mechanisms and development of resistance still not yet being fully understood^8^. For example, there is a spectrum of responses to taxane therapies among mCRPC patient populations due to variations in biological and genetic factors, but DTX resistance inevitably occurs^33^. In this study, we have shown that CLF induces significant cytotoxicity in PCa cell lines, including DTX-resistant lines, as single agent and synergistically, in combination with standard-of-care taxane treatment, which could be used as a therapeutic strategy for overcoming taxane resistance. Next, using cell-based studies and scRNAseq and RNAseq-based next-generation gene expression profiling, we have provided additional insights into the mechanism of action for CLF. Notably, we showed that CLF significantly dysregulated key non-coding RNAs associated with tumor progression and drug resistance in lethal PCa.

Earlier studies have shown that CLF functions as a broad-spectrum antibiotic by generating reactive oxygen species, superoxide, and hydrogen peroxide, as well as exhibiting anti-inflammatory properties in leprosy and autoimmune diseases^12^. While the exact mechanisms of action of CLF are under investigation, we showed that CLF activates the peroxisome proliferator-activated receptor-gamma (PPAR-γ) and synergizes with tyrosine-kinase inhibitors in the treatment of CML^31^. Additionally, we proposed several novel potential pathways dysregulated by CLF treatment in myeloma, including induction of ER stress, autophagy, mitochondrial dysfunction, oxidative phosphorylation, enhancement of the downstream cascade of p65-NFkB-IRF4-Myc downregulation, and ROS-dependent apoptotic cell death^14^. Earlier studies have shown that Clofazimine inhibits the proliferation of squamous hepatocellular carcinoma and lung cancer cell lines^34,35^. A recent study revealed that Clofazimine exerts an anti-cancer effect by inhibiting Wnt signaling in several cancer cell lines representing hepatocellular carcinoma, glioblastoma, colorectal, and ovarian cancer^36^. Further, a clinical drug screening showed Clofazimine potentiates the anti-tumor efficacy and reduces the toxicity of dual anti-PD-1 and CTLA-4 immune checkpoint blockade (ICB)^37^. CLF’s potential for the treatment of mCRPC was initially identified through an unbiased phenotype-based screen that analyzed 1,120 different compounds, where CLF showed preferential cytotoxicity and apoptotic effects for mCRPC cells, both *in vitro* and *in vivo* ^13^. As no previous studies have further explored the efficacy of CLF in PCa cells, our work has extended the knowledge on this potential therapeutic strategy, established the synergistic effects of CLF and DTX, and, by using integrated-omics-based strategies, explored the mechanism of action of CLF in mCRPC cell lines. CLF’s potential for the treatment of mCRPC was validated through *in vitro* chemosensitivity assays, where it was shown that CLF induces cytotoxicity and caspase-dependent apoptosis in PCa cell lines, both as a single agent and in combination with DTX. While the CLF doses that showed synergy were in the micromolar concentration range, they are within the safe and well-tolerated dose range of 0.84-8.4 µM, corresponding to a human C_max_ of 0.4-4 mg/L^12^. The potential synergistic effects of CLF with DTX for mCRPC cases is an avenue for further exploration through patient-derived xenograft *in vivo* models. Moreover, CLF treatment showed reduction of stem-like cell population, a key factor for EMT in PCa cell lines. For example, we showed that CLF reduces aldehyde dehydrogenase (ALDH) activity significantly in our panel of PCa cell lines compared to control cells. ALDH is a surface marker that characterizes cancer stem cells (CSCs), which are defined as malignant tumor cells with a high potential for self-renewal and differentiation^38^. CSCs derived from PCa express ALDH, and chemotherapeutic resistance can be induced by CSCs through ROS generation and ALDH activity^38,39^. Treatment of our panel of PCa cell lines with CLF reduced ALDH activity significantly in most cell lines, including a notable 67.86% decrease in the taxane-resistant cell line. Cell dissociation from the primary tumor site and migration to secondary sites through confined channels is key to cell proliferation in distant organs & cell plasticity (EMT), and cancer metastatic spread^39–41^. We designed the PDMS-based microfluidic device as a physiologically relevant model of metastasis using standard multilayer photolithography and replica molding to study the effect of our drug combination on tumor cell motility through μ-channels of dimensions that mimic the size of channel-like tracks encountered by migrating cells *in vivo*^24,42^. Our results showed that CLF significantly decreased the migration efficiency of mCRPC cells compared to control.

We have previously shown in chronic myeloid leukemia and multiple myeloma studies that CLF modulates PPARγ ubiquitin ligase activity, which results in ROS-mediated apoptosis^14,31^. PPARγ functions as a tumor suppressor in PCa that induces inhibition of cell growth, decreased tumor invasiveness, and decreased production of proinflammatory cytokines ^43^. Here, using gene expression profiling and pathway analysis, we observed that several top differentially regulated genes are related to cellular stress pathways that are modulated by PPARγ. For example, CHAC1, which is induced following unfolded protein response, showed a positive fold change of 16.42 following CLF treatment. CHAC1 modulates several pro-apoptotic transcription factors, including ATF3 and CHOP, and thus, the up-regulation of CHAC1 is connected to increased apoptotic activity^44^. DDIT4 displayed a positive fold change of 12.20 when PCa cells were treated with CLF. This protein-coding gene is associated with the mTOR signaling pathway. Overexpression of DDIT4 is induced under cellular stress and contributes to stress-induced responses, including autophagy and apoptosis^45^.

Most importantly, CLF treatment was found to modulate the expression of several non-coding RNAs, including long non-coding RNAs, snRNAs, and RNA adjuvants, that are associated with tumor cell proliferation, cell migration, invasion, and PCa drug resistance. The overexpression of MALAT1 is associated with increased cell proliferation, invasion, metastasis, evasion of apoptosis, and drug resistance through the regulation of several cell-signaling pathways^46^. Additionally, MALAT1 is overexpressed in several aggressive solid tumors, including Breast, esophageal, and lung cancer^47–50^. Up-regulated MALAT-1 contributes to bladder cancer cell migration by inducing epithelial-to-mesenchymal transition^51^. Further, MALAT-1 is also emerging as a new potential therapeutic target for CRPC since it is correlated with high Gleason score, prostate-specific antigen, and tumor stage^52^. Higher levels of NEAT1 are known to indicate poor survival in cancer patients, are associated with progression and bone metastasis in PCa patients, and reduce sensitivity to chemotherapy^53–56^. We found that MALAT1 expression was significantly downregulated (fold-change −18.06) when PCa cells were treated with CLF. Treatment with CLF was found to decrease the level of NEAT1 < −8.01 folds. Furthermore, lncRNAs have been shown to modulate rRNA synthesis factors and nucleolar integrity, which are involved in ribosome biogenesis and the regulation of ribosomal RNA (rRNA) expression^57^. For example, MALAT1 has been implicated in pre-mRNA splicing^58^. Consistently, we observed that rRNA processing was among the top downregulated canonical pathways, suggesting the effect of CLF treatment on the non-coding RNAs-mediated rRNA synthesis. Notably, using single-cell analysis, we demonstrated that MALAT1 and NEAT1 are enriched in cell subpopulations within all 4 mCRPC cell lines, including the taxane-resistant mCRPC cell line DUTXR, indicating a potential benefit of treating patients with castration-resistant and drug-resistant PCa with Clofazimine.

RTCA-AS1, which is associated with pathological grade oral and oropharyngeal squamous cell carcinoma, is decreased by a fold change of 4.28 in CLF-treated PCa cells^59^. LINC00973 is a lncRNA that is over-expressed following DNA damage and has anti-proliferative and pro-apoptotic functions through inhibition of p21 degradation^60^. LINC00973 is also up-regulated in breast cancer and promotes tumor growth by regulating MAPK signaling^61^. After CLF treatment of PCa cells, the level of LINC00973 present was increased by a fold-change of 6.55. Additionally, RN7SK is a lncRNA that has been shown to be associated with a positive feedback circuit through interaction with m6A readers in tumor cells, which increases cellular proliferation^62^. PCa cells treated with CLF downregulated the expression of RN7SK by greater than 10-fold (Fold change −10.29) compared to untreated control. STAG3L5P-PVRIG2P-PILRB is up-regulated in bladder cancer and has been identified as a prognostic necroptosis-related lncRNA^63^. Treatment with CLF decreased the expression of STAG3L5P-PVRIG2P-PILRB (Fold-change −3.85) in mCRPC cells. LOXL-AS1 is known to regulate gene expression and signaling pathways through binding to microRNAs, affecting cell proliferation, migration, invasion, EMT, and apoptosis^64,65^. It is also up-regulated in multiple cancers, including PCa, and is positively associated with adverse clinical signs in cancer patients^64,66^. LOXL-AS1 expression was decreased in CLF-treated PCa cells by ∼3 folds (Fold change −2.90). LINC00342 has shown a correlation between its overexpression and poor prognosis of patients with clear cell renal cell carcinoma, likely due to its role in the reprogramming of glucose metabolism and cancer metastasis^67^.

Finally, the non-coding RNA 7SL is highly expressed in cancer tissues and involved in the post-transcriptional regulation of several important genes, including repression of translation of the tumor suppressor p53 by interacting with the 3′UTR of TP53 mRNA^68^. This mechanism has been shown to mediate the proliferative effects of 7SL, resulting in repression of p53 translation and the accumulation of p53 in cancer cells. 7SL is encoded by RN7SL1 (RNA component of the signal recognition particle 7SL1) and possesses a small ORF (smORF) that has been shown to be translated into peptides in cancer^69^. RN7SL2 is another small, cytoplasmic RNA molecule that is highly expressed in the cell-free plasma of cancer patients^70^. Our analysis of gene expression on clinical datasets showed that high expression of RN7SL1 and RN7SL2 was associated with significant biochemical recurrence. Since we observed that CLF treatment significantly downregulated these non-coding RNAs, this further indicates that CLF treatment may be clinically effective in treating these patients with advanced/aggressive PCa.

Since CLF is an FDA-approved low-cost medication that is well-tolerated in patients and also on the World Health Organization’s List of Essential Medicines, CLF is an exciting potential approach for the treatment of highly aggressive and drug-resistant mCRPC.

## Authorship Contributions

S.B., H.K., T.M.G., and A.K.M. participated in the research design; S.B., H.K., D.C.N., S.M., and T.M.G. conducted experiments; S.B., H.K., P.M., T.M.G., and A.K.M. performed data analysis; S.B., P.M., R.A., T.M.G., and A.K.M. wrote or contributed to the writing of the manuscript; A.K.M. supervised the overall project. All authors have read and agreed to the published version of the manuscript.

## Data Availability

The datasets generated during and/or analyzed during the current study are available from the corresponding author upon reasonable request.

