## Supplementary Material for "Clofazimine treatment modulates key non-coding RNAs associated with tumor progression and drug resistance in lethal Prostate Cancer"

Sarah Batten *et al*

**This file includes:**

Supplementary Figure. S1
Supplementary Tables S1 to S3

**Supplementary Figure S1.** Single-agent in vitro cytotoxicity of Clofazimine in mCRPC cell lines.

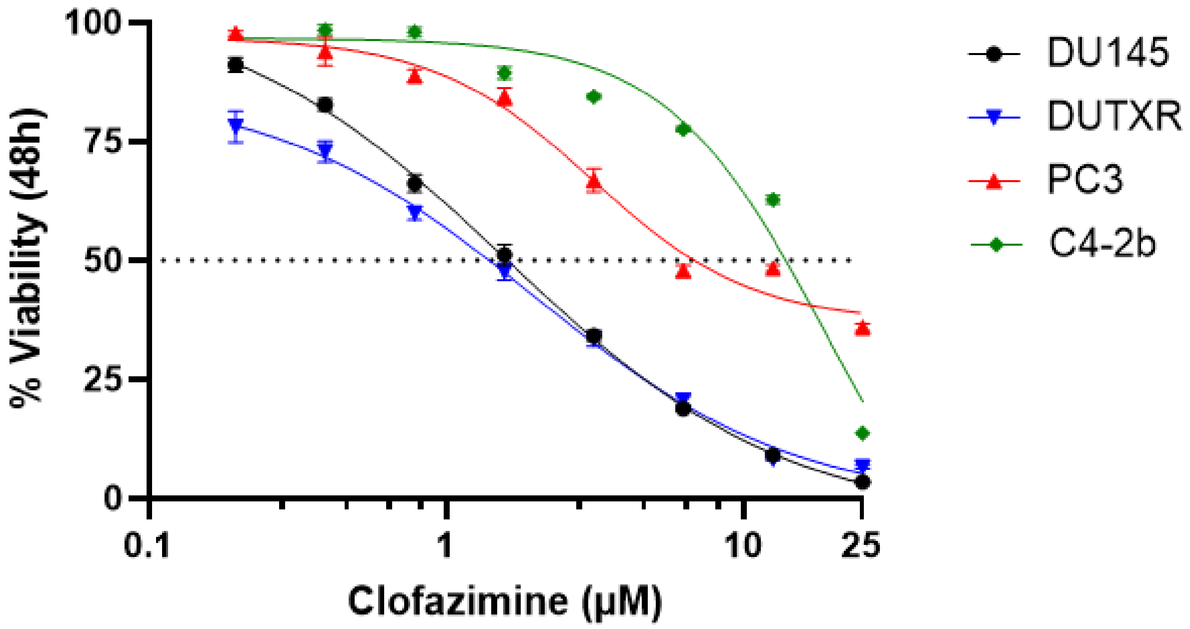


| | DU145 | DUTXR | PC3 | C4-2b |
|---|---|---|---|---|
| CLF (μM) | 1.59 | 1.965 | 3.095 | 18.81 |
| DTX (nM) | 5.547 | 962.3 | 3.377 | 1.416 |

**Supplementary Table S1.** List of drugs and reagents used in the study.

| Reagents | Manufacturer | Location |
|---|---|---|
| Fetal bovine serum | Hyclone (Thermo-Fisher Scientific Inc.) | Rockford, IL, USA |
| Trypsin (0.25% w/v) | Hyclone (Thermo-Fisher Scientific Inc.) | Rockford, IL, USA |
| Penicillin-Streptomycin (10,000 U/mL) | Gibco™ (Thermo-Fisher Scientific Inc.) | Waltham, MA, USA |
| FITC Annexin V Apoptosis Detection Kit | BD Biosciences | San Jose, CA, USA |
| RIPA lysis buffer | Thermo-Fisher Scientific Inc. | Waltham, MA, USA |
| Halt™ Protease and Phosphatase Inhibitor Cocktail | Thermo-Fisher Scientific Inc. | Waltham, MA, USA |
| Pierce ™ ECL Western Blotting Substrate | Thermo-Fisher Scientific Inc. | Waltham, MA, USA |
| RNeasy Plus Mini Kit | QIAGEN | Hilden, Germany |
| Quick Start Bovine Serum Albumin Standard | Bio-Rad | Hercules, CA, USA |
| Tris Buffer Saline (TBS) | Bio-Rad | Hercules, CA, USA |
| 10% Tween 20 | Bio-Rad | Hercules, CA, USA |
| Polyvinylidene fluoride membrane (PVDF) | EMD Millipore | Billerica, MA, USA |
| Bovine Serum Albumin (BSA) | VWR | Radnor, PA, USA |
| Dimethyl sulfoxide (DMSO) | Sigma-Aldrich Inc. | St. Louis, MO, USA |
| Bradford Reagent | Sigma-Aldrich Inc. | St. Louis, MO, USA |
| JC-1 - Mitochondrial Membrane Potential Assay Kit | Abcam | Waltham, MA, USA |
| Anti-rabbit IgG, HRP-linked Antibody (7074S) | Cell Signaling Technology | Danvers, MA, USA |
| β-actin (A3854) Antibody | Sigma-Aldrich Inc | St. Louis, MO, USA |
| Clofazimine | Selleckchem | Houston, TX, USA |
| Docetaxel | Selleckchem | Houston, TX, USA |

**Supplementary Table S2**. The top coding genes in mCRPC cell lines ((FoldChange (CLF vs treatment) >|3.5|; p<0.05))

| Gene ID | Gene name | FDR step up (CLF vs No treatment) | Fold change (CLF vs No treatment) |
|---|---|---|---|
| ENSG00000228253 | **MT-ATP8** | 2.55E-63 | -62.127 |
| ENSG00000269955 | **FMC1-LUC7L2** | 2.93E-06 | -20.572 |
| ENSG00000175265 | **GOLGA8A** | 8.05E-19 | -12.901 |
| ENSG00000215252 | **GOLGA8B** | 1.90E-17 | -8.294 |
| ENSG00000187244 | **BCAM** | 2.23E-18 | -8.056 |
| ENSG00000131746 | **TNS4** | 2.96E-16 | -7.755 |
| ENSG00000126709 | **IFI6** | 1.09E-16 | -7.362 |
| ENSG00000117472 | **TSPAN1** | 1.07E-13 | -7.330 |
| ENSG00000198763 | **MT-ND2** | 0.00E+00 | -5.971 |
| ENSG00000120738 | **EGR1** | 3.50E-03 | -5.597 |
| ENSG00000198786 | **MT-ND5** | 0.00E+00 | -5.477 |
| ENSG00000198888 | **MT-ND1** | 0.00E+00 | -5.394 |
| ENSG00000108848 | **LUC7L3** | 1.32E-33 | -5.233 |
| ENSG00000142089 | **IFITM3** | 3.09E-05 | -4.910 |
| ENSG00000065618 | **COL17A1** | 6.57E-04 | -4.722 |
| ENSG00000186919 | **ZACN** | 2.43E-02 | -4.715 |
| ENSG00000183336 | **BOLA2** | 6.25E-07 | -4.692 |
| ENSG00000198695 | **MT-ND6** | 3.36E-254 | -4.687 |
| ENSG00000135976 | **ANKRD36** | 7.69E-04 | -4.659 |
| ENSG00000102901 | **CENPT** | 1.71E-10 | -4.505 |
| ENSG00000155657 | **TTN** | 1.22E-02 | -4.225 |
| ENSG00000221978 | **CCNL2** | 4.19E-23 | -4.163 |
| ENSG00000161996 | **WDR90** | 8.89E-05 | -4.075 |
| ENSG00000158062 | **UBXN11** | 9.54E-04 | -4.004 |
| ENSG00000178038 | **ALS2CL** | 1.08E-04 | -3.954 |
| ENSG00000137070 | **IL11RA** | 1.46E-02 | -3.937 |
| ENSG00000242110 | **AMACR** | 1.20E-09 | -3.929 |
| ENSG00000198064 | **NPIPB13** | 2.31E-04 | -3.858 |
| ENSG00000212907 | **MT-ND4L** | 4.53E-234 | -3.789 |
| ENSG00000111331 | **OAS3** | 1.03E-14 | -3.736 |
| ENSG00000176155 | **CCDC57** | 3.57E-06 | -3.734 |
| ENSG00000182272 | **B4GALNT4** | 6.15E-03 | -3.696 |
| ENSG00000080493 | **SLC4A4** | 2.10E-02 | -3.657 |
| ENSG00000124664 | **SPDEF** | 2.46E-03 | -3.596 |
| ENSG00000105486 | **LIG1** | 4.34E-04 | -3.592 |
| ENSG00000127415 | **IDUA** | 4.30E-04 | -3.581 |
| ENSG00000090581 | **GNPTG** | 2.70E-10 | -3.512 |
| ENSG00000101670 | **LIPG** | 5.24E-04 | 3.559 |
| ENSG00000184897 | **H1-10** | 1.46E-57 | 3.567 |
| ENSG00000170454 | **KRT75** | 1.62E-02 | 3.568 |
| ENSG00000150907 | **FOXO1** | 2.47E-06 | 3.592 |
| ENSG00000260001 | **TGFBR3L** | 9.72E-04 | 3.603 |
| ENSG00000291237 | **SOD2** | 4.84E-114 | 3.624 |
| ENSG00000099625 | **CBARP** | 2.04E-07 | 3.663 |

| | | | |
|---|---|---|---|
| ENSG00000144426 | **NBEAL1** | 8.01E-136 | 3.744 |
| ENSG00000041982 | **TNC** | 3.85E-92 | 3.767 |
| ENSG00000154217 | **PITPNC1** | 1.21E-14 | 3.839 |
| ENSG00000128564 | **VGF** | 3.90E-05 | 3.870 |
| ENSG00000149948 | **HMGA2** | 1.30E-18 | 3.904 |
| ENSG00000175155 | **YPEL2** | 5.79E-03 | 3.919 |
| ENSG00000172159 | **FRMD3** | 2.18E-04 | 3.933 |
| ENSG00000145685 | **LHFPL2** | 1.89E-16 | 3.937 |
| ENSG00000110675 | **ELMOD1** | 7.82E-03 | 3.941 |
| ENSG00000146278 | **PNRC1** | 6.34E-29 | 3.947 |
| ENSG00000118689 | **FOXO3** | 7.94E-65 | 3.949 |
| ENSG00000043591 | **ADRB1** | 4.13E-03 | 3.957 |
| ENSG00000175556 | **LONRF3** | 7.39E-09 | 3.971 |
| ENSG00000258102 | **MAP1LC3B2** | 2.11E-14 | 3.979 |
| ENSG00000139112 | **GABARAPL1** | 2.85E-11 | 4.158 |
| ENSG00000112182 | **BACH2** | 2.40E-03 | 4.171 |
| ENSG00000120278 | **PLEKHG1** | 3.18E-02 | 4.205 |
| ENSG00000115844 | **DLX2** | 1.15E-02 | 4.217 |
| ENSG00000165312 | **OTUD1** | 4.71E-07 | 4.220 |
| ENSG00000127530 | **OR7C1** | 4.02E-06 | 4.263 |
| ENSG00000110492 | **MDK** | 1.51E-15 | 4.275 |
| ENSG00000107954 | **NEURL1** | 4.16E-02 | 4.291 |
| ENSG00000167157 | **PRRX2** | 2.71E-03 | 4.298 |
| ENSG00000137285 | **TUBB2B** | 4.21E-04 | 4.314 |
| ENSG00000069667 | **RORA** | 8.88E-05 | 4.377 |
| ENSG00000245848 | **CEBPA** | 7.94E-06 | 4.443 |
| ENSG00000134070 | **IRAK2** | 4.55E-19 | 4.461 |
| ENSG00000155961 | **RAB39B** | 1.59E-02 | 4.634 |
| ENSG00000123610 | **TNFAIP6** | 4.21E-03 | 4.760 |
| ENSG00000187479 | **C11orf96** | 4.02E-02 | 4.774 |
| ENSG00000175197 | **DDIT3** | 3.56E-22 | 4.806 |
| ENSG00000163661 | **PTX3** | 3.10E-02 | 4.806 |
| ENSG00000025039 | **RRAGD** | 5.96E-06 | 4.846 |
| ENSG00000131459 | **GFPT2** | 4.73E-09 | 5.125 |
| ENSG00000154133 | **ROBO4** | 1.46E-03 | 5.133 |
| ENSG00000115602 | **IL1RL1** | 7.70E-04 | 5.255 |
| ENSG00000100292 | **HMOX1** | 3.82E-20 | 5.347 |
| ENSG00000176170 | **SPHK1** | 3.73E-20 | 5.418 |
| ENSG00000136842 | **TMOD1** | 3.85E-03 | 5.451 |
| ENSG00000145107 | **TM4SF19** | 5.09E-05 | 5.502 |
| ENSG00000104369 | **JPH1** | 1.75E-08 | 5.536 |
| ENSG00000168398 | **BDKRB2** | 7.66E-03 | 5.623 |
| ENSG00000221869 | **CEBPD** | 1.43E-12 | 5.680 |
| ENSG00000115919 | **KYNU** | 1.29E-03 | 5.711 |
| ENSG00000161011 | **SQSTM1** | 0.00E+00 | 5.760 |
| ENSG00000008517 | **IL32** | 3.36E-15 | 5.931 |
| ENSG00000128965 | **CHAC1** | 4.18E-14 | 6.085 |
| ENSG00000165521 | **EML5** | 4.92E-12 | 6.113 |
| ENSG00000134363 | **FST** | 1.39E-10 | 6.126 |

| | | | |
|---|---|---|---|
| ENSG00000168209 | **DDIT4** | 3.13E-65 | 6.519 |
| ENSG00000161681 | **SHANK1** | 1.35E-05 | 6.674 |
| ENSG00000138166 | **DUSP5** | 5.73E-108 | 7.044 |
| ENSG00000107249 | **GLIS3** | 1.10E-12 | 7.138 |
| ENSG00000049249 | **TNFRSF9** | 8.08E-04 | 8.786 |
| ENSG00000073756 | **PTGS2** | 6.62E-04 | 9.076 |
| ENSG00000124102 | **PI3** | 1.16E-08 | 10.307 |
| ENSG00000125730 | **C3** | 5.71E-08 | 10.325 |
| ENSG00000169429 | **CXCL8** | 6.51E-28 | 16.053 |
| ENSG00000167995 | **BEST1** | 3.45E-38 | 18.396 |
| ENSG00000148346 | **LCN2** | 5.13E-50 | 53.300 |

**Supplementary Table S3**. Top differentially regulated genes following CLF treatment in DUTXR cell line **(FoldChange (CLF vs treatment) >|3.5|; p<0.05)**

| Gene ID | Gene name | Gene biotype | FDR step up (CLF vs No treatment) | Fold change (CLF vs No treatment) |
|---|---|---|---|---|
| ENSG00000120738 | **EGR1** | protein_coding | 2.6E-19 | -24.04 |
| ENSG00000168743 | **NPNT** | protein_coding | 5.0E-07 | -23.28 |
| ENSG00000250722 | **SELENOP** | protein_coding | 7.2E-03 | -17.09 |
| ENSG00000115461 | **IGFBP5** | protein_coding | 1.7E-03 | -16.32 |
| ENSG00000265972 | **TXNIP** | protein_coding | 8.5E-17 | -16.27 |
| ENSG00000124406 | **ATP8A1** | protein_coding | 5.9E-07 | -16.06 |
| ENSG00000170345 | **FOS** | protein_coding | 9.3E-10 | -15.41 |
| ENSG00000276168 | **RN7SL1** | misc_RNA | 1.6E-58 | -12.63 |
| ENSG00000172986 | **GXYLT2** | protein_coding | 1.3E-02 | -11.04 |
| ENSG00000165023 | **DIRAS2** | protein_coding | 3.6E-02 | -9.32 |
| ENSG00000168646 | **AXIN2** | protein_coding | 9.0E-04 | -9.23 |
| ENSG00000151632 | **AKR1C2** | protein_coding | 4.2E-02 | -9.12 |
| ENSG00000106772 | **PRUNE2** | protein_coding | 1.2E-10 | -8.71 |
| ENSG00000119699 | **TGFB3** | protein_coding | 7.6E-03 | -8.68 |
| ENSG00000164938 | **TP53INP1** | protein_coding | 5.9E-04 | -8.65 |
| ENSG00000274070 | **CASTOR2** | protein_coding | 9.6E-06 | -8.23 |
| ENSG00000196139 | **AKR1C3** | protein_coding | 3.6E-09 | -8.02 |
| ENSG00000180596 | **H2BC4** | protein_coding | 2.8E-04 | -7.81 |
| ENSG00000106780 | **MEGF9** | protein_coding | 2.3E-61 | -7.71 |
| ENSG00000078018 | **MAP2** | protein_coding | 3.5E-08 | -7.41 |
| ENSG00000274012 | **RN7SL2** | misc_RNA | 3.7E-49 | -6.94 |
| ENSG00000149212 | **SESN3** | protein_coding | 3.2E-36 | -6.84 |
| ENSG00000138449 | **SLC40A1** | protein_coding | 1.8E-03 | -6.81 |
| ENSG00000071967 | **CYBRD1** | protein_coding | 3.0E-32 | -6.63 |
| ENSG00000103888 | **CEMIP** | protein_coding | 1.9E-04 | -6.63 |
| ENSG00000171903 | **CYP4F11** | protein_coding | 1.5E-02 | -6.28 |
| ENSG00000256043 | **CTSO** | protein_coding | 1.4E-02 | -6.27 |
| ENSG00000283526 | **PRRT1B** | protein_coding | 1.6E-02 | -6.21 |
| ENSG00000234664 | **HMGN2P5** | processed_pseudogene | 8.2E-03 | -6.18 |
| ENSG00000167703 | **SLC43A2** | protein_coding | 9.2E-07 | -6.16 |
| ENSG00000163683 | **SMIM14** | protein_coding | 9.3E-30 | -5.89 |
| ENSG00000145358 | **DDIT4L** | protein_coding | 1.5E-02 | -5.88 |
| ENSG00000047644 | **WWC3** | protein_coding | 2.3E-10 | -5.80 |
| ENSG00000188042 | **ARL4C** | protein_coding | 1.5E-07 | -5.75 |
| ENSG00000170962 | **PDGFD** | protein_coding | 1.2E-04 | -5.66 |
| ENSG00000111077 | **TNS2** | protein_coding | 1.1E-04 | -5.59 |
| ENSG00000119711 | **ALDH6A1** | protein_coding | 4.3E-03 | -5.58 |
| ENSG00000134962 | **KLB** | protein_coding | 8.6E-04 | -5.54 |
| ENSG00000129595 | **EPB41L4A** | protein_coding | 2.6E-02 | -5.47 |

| | | | | |
|---|---|---|---|---|
| ENSG00000114315 | **HES1** | protein_coding | 9.4E-12 | -5.38 |
| ENSG00000228253 | **MT-ATP8** | protein_coding | 2.9E-04 | -5.37 |
| ENSG00000166147 | **FBN1** | protein_coding | 6.9E-19 | -5.27 |
| ENSG00000143126 | **CELSR2** | protein_coding | 2.0E-12 | -5.20 |
| ENSG00000204396 | **VWA7** | protein_coding | 4.6E-03 | -5.17 |
| ENSG00000125966 | **MMP24** | protein_coding | 8.6E-10 | -5.13 |
| ENSG00000106789 | **CORO2A** | protein_coding | 3.6E-03 | -5.13 |
| ENSG00000114698 | **PLSCR4** | protein_coding | 5.5E-03 | -5.06 |
| ENSG00000169962 | **TAS1R3** | protein_coding | 4.6E-03 | -5.06 |
| ENSG00000156298 | **TSPAN7** | protein_coding | 3.0E-07 | -4.98 |
| ENSG00000076864 | **RAP1GAP** | protein_coding | 3.4E-06 | -4.94 |
| ENSG00000149131 | **SERPING1** | protein_coding | 7.3E-05 | -4.88 |
| ENSG00000121005 | **CRISPLD1** | protein_coding | 6.4E-07 | -4.87 |
| ENSG00000080493 | **SLC4A4** | protein_coding | 2.6E-05 | -4.72 |
| ENSG00000184678 | **H2BC21** | protein_coding | 4.9E-06 | -4.70 |
| ENSG00000136040 | **PLXNC1** | protein_coding | 8.3E-03 | -4.69 |
| ENSG00000196730 | **DAPK1** | protein_coding | 1.5E-04 | -4.63 |
| ENSG00000172037 | **LAMB2** | protein_coding | 2.9E-43 | -4.61 |
| ENSG00000108375 | **RNF43** | protein_coding | 2.6E-06 | -4.58 |
| ENSG00000278771 | **RN7SL3** | misc_RNA | 1.6E-02 | -4.56 |
| ENSG00000126709 | **IFI6** | protein_coding | 2.6E-07 | -4.53 |
| ENSG00000183336 | **BOLA2** | protein_coding | 1.5E-03 | -4.51 |
| ENSG00000138798 | **EGF** | protein_coding | 5.7E-04 | -4.51 |
| ENSG00000126016 | **AMOT** | protein_coding | 3.8E-03 | -4.47 |
| ENSG00000211459 | **MT-RNR1** | Mt_rRNA | 4.5E-55 | -4.43 |
| ENSG00000118898 | **PPL** | protein_coding | 1.3E-10 | -4.42 |
| ENSG00000182580 | **EPHB3** | protein_coding | 1.8E-04 | -4.37 |
| ENSG00000205213 | **LGR4** | protein_coding | 5.7E-21 | -4.36 |
| ENSG00000146233 | **CYP39A1** | protein_coding | 4.0E-02 | -4.34 |
| ENSG00000185745 | **IFIT1** | protein_coding | 1.2E-03 | -4.30 |
| ENSG00000164300 | **SERINC5** | protein_coding | 4.9E-08 | -4.26 |
| ENSG00000198513 | **ATL1** | protein_coding | 3.3E-02 | -4.20 |
| ENSG00000173376 | **NDNF** | protein_coding | 3.3E-02 | -4.19 |
| ENSG00000291122 | **CASTOR3P** | lncRNA | 3.5E-03 | -4.19 |
| ENSG00000121691 | **CAT** | protein_coding | 3.6E-12 | -4.17 |
| ENSG00000187134 | **AKR1C1** | protein_coding | 4.5E-04 | -4.13 |
| ENSG00000185585 | **OLFML2A** | protein_coding | 8.0E-03 | -4.11 |
| ENSG00000006534 | **ALDH3B1** | protein_coding | 7.4E-08 | -4.07 |
| ENSG00000143512 | **HHIPL2** | protein_coding | 1.3E-03 | -3.98 |
| ENSG00000149809 | **TM7SF2** | protein_coding | 1.2E-05 | -3.98 |
| ENSG00000173698 | **ADGRG2** | protein_coding | 1.6E-05 | -3.95 |
| ENSG00000251562 | **MALAT1** | lncRNA | 1.2E-26 | -3.90 |
| ENSG00000181524 | **RPL24P4** | processed_pseudogene | 1.7E-08 | -3.85 |

| | | | | |
|---|---|---|---|---|
| ENSG00000118785 | **SPP1** | protein_coding | 1.6E-03 | -3.80 |
| ENSG00000127415 | **IDUA** | protein_coding | 2.0E-02 | -3.79 |
| ENSG00000108679 | **LGALS3BP** | protein_coding | 1.3E-09 | -3.75 |
| ENSG00000124171 | **PARD6B** | protein_coding | 3.0E-04 | -3.69 |
| ENSG00000128833 | **MYO5C** | protein_coding | 2.6E-06 | -3.66 |
| ENSG00000123358 | **NR4A1** | protein_coding | 7.5E-09 | -3.61 |
| ENSG00000165757 | **JCAD** | protein_coding | 1.0E-02 | -3.55 |
| ENSG00000219507 | **FTH1P8** | processed_pseudogene | 1.7E-04 | -3.54 |
| ENSG00000169220 | **RGS14** | protein_coding | 4.8E-02 | -3.53 |
| ENSG00000186951 | **PPARA** | protein_coding | 5.9E-03 | -3.53 |
| ENSG00000163584 | **RPL22L1** | protein_coding | 8.8E-03 | 3.50 |
| ENSG00000070669 | **ASNS** | protein_coding | 8.7E-08 | 3.50 |
| ENSG00000177954 | **RPS27** | protein_coding | 7.5E-30 | 3.52 |
| ENSG00000253368 | **TRNP1** | protein_coding | 2.5E-67 | 3.62 |
| ENSG00000169715 | **MT1E** | protein_coding | 5.3E-04 | 3.63 |
| ENSG00000232956 | **SNHG15** | lncRNA | 4.2E-05 | 3.65 |
| ENSG00000101255 | **TRIB3** | protein_coding | 2.7E-14 | 3.66 |
| ENSG00000117143 | **UAP1** | protein_coding | 2.1E-15 | 3.68 |
| ENSG00000172780 | **RAB43** | protein_coding | 1.6E-02 | 3.69 |
| ENSG00000162496 | **DHRS3** | protein_coding | 5.4E-03 | 3.71 |
| ENSG00000139289 | **PHLDA1** | protein_coding | 5.2E-29 | 3.71 |
| ENSG00000163347 | **CLDN1** | protein_coding | 2.5E-10 | 3.72 |
| ENSG00000175197 | **DDIT3** | protein_coding | 7.8E-04 | 3.76 |
| ENSG00000159167 | **STC1** | protein_coding | 4.4E-02 | 3.83 |
| ENSG00000164509 | **IL31RA** | protein_coding | 2.6E-03 | 3.84 |
| ENSG00000196756 | **SNHG17** | lncRNA | 4.5E-05 | 3.95 |
| ENSG00000244716 | **RPL17P7** | processed_pseudogene | 4.8E-12 | 3.97 |
| ENSG00000103044 | **HAS3** | protein_coding | 3.1E-05 | 3.98 |
| ENSG00000160193 | **WDR4** | protein_coding | 1.3E-03 | 3.99 |
| ENSG00000011422 | **PLAUR** | protein_coding | 4.0E-16 | 4.01 |
| ENSG00000049249 | **TNFRSF9** | protein_coding | 4.8E-02 | 4.10 |
| ENSG00000251493 | **FOXD1** | protein_coding | 1.2E-12 | 4.10 |
| ENSG00000026508 | **CD44** | protein_coding | 1.1E-37 | 4.12 |
| ENSG00000164647 | **STEAP1** | protein_coding | 6.0E-05 | 4.16 |
| ENSG00000179431 | **FJX1** | protein_coding | 5.0E-05 | 4.21 |
| ENSG00000148848 | **ADAM12** | protein_coding | 4.2E-02 | 4.30 |
| ENSG00000128965 | **CHAC1** | protein_coding | 8.5E-03 | 4.34 |
| ENSG00000186854 | **TRABD2A** | protein_coding | 9.9E-03 | 4.36 |
| ENSG00000058085 | **LAMC2** | protein_coding | 2.4E-09 | 4.44 |
| ENSG00000206190 | **ATP10A** | protein_coding | 3.6E-03 | 4.50 |
| ENSG00000213261 | **EEF1B2P6** | processed_pseudogene | 2.3E-02 | 4.51 |
| ENSG00000185022 | **MAFF** | protein_coding | 2.8E-04 | 4.57 |
| ENSG00000139278 | **GLIPR1** | protein_coding | 5.9E-08 | 4.60 |

| | | | | |
|---|---|---|---|---|
| ENSG00000138166 | **DUSP5** | protein_coding | 2.3E-07 | 4.72 |
| ENSG00000187534 | **PRR13P5** | processed_pseudogene | 3.3E-02 | 4.77 |
| ENSG00000134531 | **EMP1** | protein_coding | 6.9E-04 | 4.79 |
| ENSG00000178464 | **RPL10P16** | processed_pseudogene | 1.8E-11 | 4.90 |
| ENSG00000188766 | **SPRED3** | protein_coding | 7.1E-04 | 4.94 |
| ENSG00000118503 | **TNFAIP3** | protein_coding | 8.9E-03 | 5.06 |
| ENSG00000135480 | **KRT7** | protein_coding | 3.7E-15 | 5.18 |
| ENSG00000141526 | **SLC16A3** | protein_coding | 9.5E-07 | 5.22 |
| ENSG00000198918 | **RPL39** | protein_coding | 1.3E-09 | 5.25 |
| ENSG00000175592 | **FOSL1** | protein_coding | 2.9E-17 | 5.26 |
| ENSG00000129226 | **CD68** | protein_coding | 2.0E-07 | 5.51 |
| ENSG00000147872 | **PLIN2** | protein_coding | 1.2E-03 | 5.52 |
| ENSG00000183696 | **UPP1** | protein_coding | 3.9E-12 | 5.64 |
| ENSG00000154217 | **PITPNC1** | protein_coding | 1.2E-02 | 5.64 |
| ENSG00000101187 | **SLCO4A1** | protein_coding | 2.6E-24 | 5.83 |
| ENSG00000148677 | **ANKRD1** | protein_coding | 1.6E-24 | 5.88 |
| ENSG00000041982 | **TNC** | protein_coding | 3.1E-28 | 5.97 |
| ENSG00000139211 | **AMIGO2** | protein_coding | 3.0E-04 | 5.99 |
| ENSG00000175764 | **TTLL11** | protein_coding | 3.2E-03 | 6.18 |
| ENSG00000227097 | **RPS28P7** | processed_pseudogene | 4.9E-02 | 6.40 |
| ENSG00000154127 | **UBASH3B** | protein_coding | 8.9E-21 | 6.45 |
| ENSG00000163735 | **CXCL5** | protein_coding | 6.4E-03 | 6.54 |
| ENSG00000128283 | **CDC42EP1** | protein_coding | 4.1E-10 | 6.87 |
| ENSG00000135318 | **NT5E** | protein_coding | 4.4E-10 | 6.94 |
| ENSG00000163661 | **PTX3** | protein_coding | 6.9E-03 | 7.03 |
| ENSG00000175832 | **ETV4** | protein_coding | 1.9E-06 | 7.26 |
| ENSG00000074416 | **MGLL** | protein_coding | 3.4E-14 | 7.67 |
| ENSG00000169627 | **BOLA2B** | protein_coding | 5.9E-07 | 7.72 |
| ENSG00000136167 | **LCP1** | protein_coding | 4.7E-03 | 8.20 |
| ENSG00000197467 | **COL13A1** | protein_coding | 1.0E-02 | 8.73 |
| ENSG00000069482 | **GAL** | protein_coding | 2.0E-10 | 8.94 |
| ENSG00000090339 | **ICAM1** | protein_coding | 5.6E-06 | 9.08 |
| ENSG00000113083 | **LOX** | protein_coding | 1.2E-07 | 9.14 |
| ENSG00000163395 | **IGFN1** | protein_coding | 5.8E-05 | 9.24 |
| ENSG00000070182 | **SPTB** | protein_coding | 3.5E-02 | 9.94 |
| ENSG00000134668 | **SPOCD1** | protein_coding | 7.0E-07 | 11.07 |
| ENSG00000223617 | **LINC00370** | lncRNA | 1.4E-05 | 12.58 |
| ENSG00000183691 | **NOG** | protein_coding | 3.4E-05 | 14.02 |
| ENSG00000249992 | **TMEM158** | protein_coding | 1.7E-12 | 15.55 |
| ENSG00000176170 | **SPHK1** | protein_coding | 3.0E-06 | 16.26 |
| ENSG00000144583 | **MARCHF4** | protein_coding | 4.7E-04 | 26.93 |
